# Genetic impairment of ketone body signalling is a prevalent contributor to human metabolic dysfunction

**DOI:** 10.1101/2025.09.30.25336995

**Authors:** Alexandra R. Yesian, Brian Y. H. Lam, Hye In Kim, Felix R. Day, Alice Williamson, Raina Jia, Sam Lockhart, Kara Rainbow, Vasileios Kaimakis, Mairi Antypa, Vladimir Saudek, Julia Jones, Claire Normand, Meriem Semache, Laurent Sabbagh, Matthew J. Neville, David Araújo-Vilar, Isabelle Jéru, Kimberly A. Stevens, Jimmy X. Kong, Mitchell E. Granade, Nalissa Amar, Michael Mazzocca, Kevin M. Tveter, Joanne M. Buxton, Larry C. James, Ken K. Ong, John A. Tadross, Fredrik Karpe, David B. Savage, Daniel J. Fazakerley, Nick Wareham, John R. B. Perry, Kendra K. Bence, Jean-Philippe Fortin, Stephen O’Rahilly, Xiaoming Liu

**Author notes:** Joint first authors (Alexandra R. Yesian, Brian Y. H. Lam). Joint second authors (Hye In Kim, Felix R. Day). Joint correspondence authors.

## Abstract

Emerging evidence that circulating levels of key metabolic intermediates are sensed by a range of G-Protein Coupled receptors (GPCRs) is providing critical new insights into the control of systemic metabolic homeostasis, and how disturbances in such sensing may contribute to metabolic disease. The hydroxycarboxylic acid receptors for lactate (HCAR1), β-hydroxybutyrate (HCAR2), and octanoate (HCAR3) are encoded by three closely homologous GPCR genes co-located in a region where common genetic variation has been reportedly associated with lipid levels and body fat distribution. By resolving sequence homology in this region, we were able to refine this signal to a coding variant (R311C) in HCAR2. Using corrected genotypes from ∼500K participants from UK Biobank and direct genotyping of four other studies, we found that carriage of the *HCAR2 p.R311C* variant was significantly associated with type 2 diabetes risk, reduced gynoid fat mass, increased waist-hip ratio, higher circulating triglycerides, glucose and alanine aminotransferase levels, lower levels of HDL cholesterol and adiponectin and impaired suppression of circulating levels of non-esterified fatty acids after oral glucose. Adipose tissue explants from mice engineered to express the equivalent mutation variant (p.R308C) in the mouse ortholog showed increased lipolytic activity, basally and after β-hydroxybutyrate (BHB) treatment. In vivo, the mice were insulin resistant and had increased liver fat and impaired post-prandial suppression of NEFAs. The variant alters an amino acid located in the intracellular C-terminal tail of HCAR2, increasing recruitment of β-arrestin and resulting in enhanced internalisation and reduced cell surface expression. In conclusion, a common variant in the human ketone body receptor results in impaired control of adipocyte lipolysis and adversely impacts systemic lipid and glucose metabolism. These findings highlight the importance of anti-lipolytic ketone body signalling in adipocytes for the maintenance of metabolic health

**Graphical Abstract:** 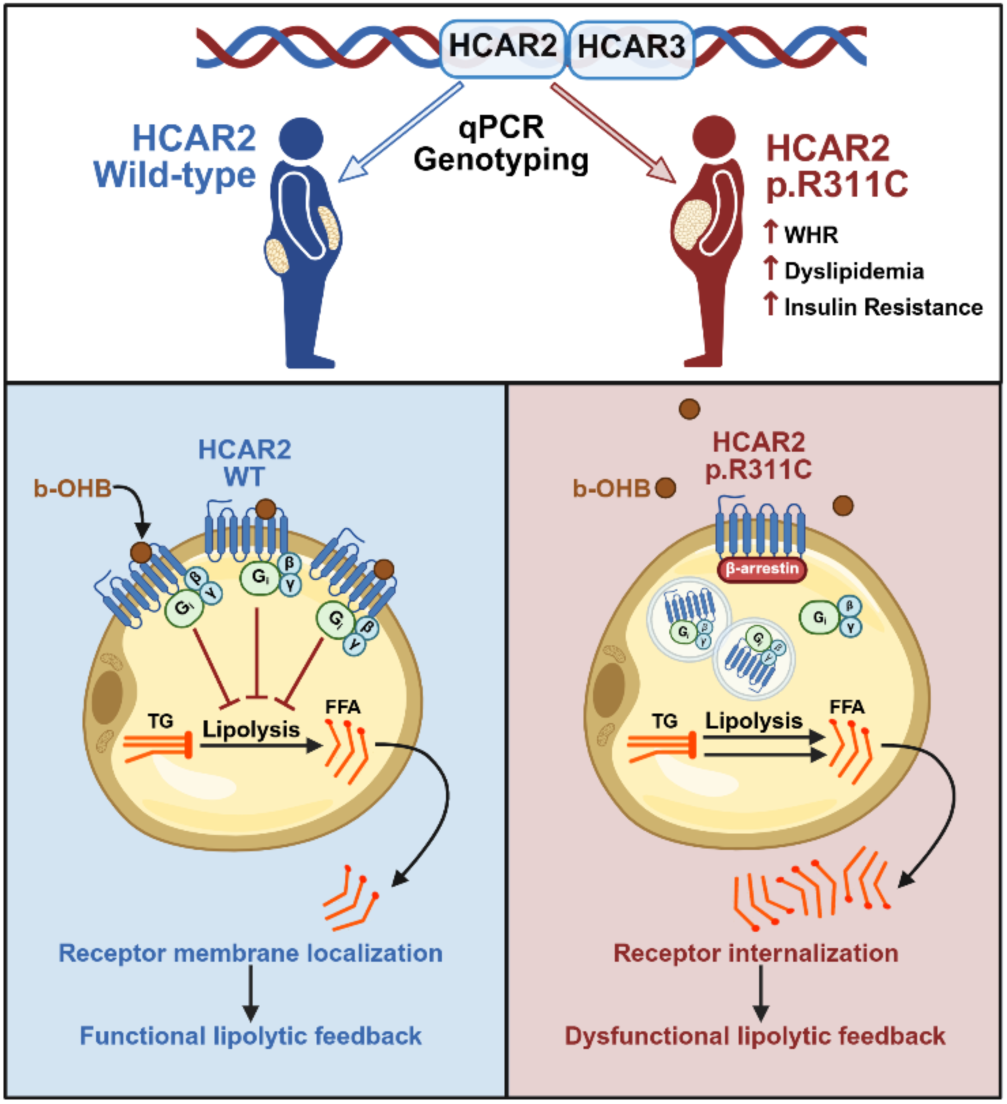

## Introduction

Central adiposity, insulin resistance, and dyslipidaemia commonly co-exist as key components of a “metabolic syndrome” that strongly predisposes to cardiovascular disease, liver disease, and Type 2 diabetes^1^. Genome wide studies of both common and rare genetic variants predisposing to individual and collective components of the metabolic syndrome have highlighted a striking enrichment of genes expressed in fat cells^2–6^.

Healthy adipocytes efficiently store excess dietary calories as triglyceride within large intracellular lipid droplets and rapidly release free fatty acids and glycerol in response to systemic demands^7^. Understanding how genetic risk variants for the metabolic syndrome disturb adipocyte function is a key goal of human metabolic research. Such predisposing variants could potentially influence a broad range of cellular processes, including adipocyte differentiation, proliferation and survival, and the control of lipogenesis and lipolysis. At least two monogenic forms of human lipodystrophy (PLIN1^8^, CIDEC^9^) result from a primary inability of adipocytes to adequately control lipolysis; the unregulated release of free fatty acids and glycerol results in lipotoxic effects on other organs, such as muscle and liver. Several rare genetic variants influencing metabolic syndrome risk also appear to converge on the lipolytic process, including receptor systems that respond to circulating regulators of lipolysis^3,10^.

In addition to responding to hormones, adipocytes also express receptors for certain key metabolites such as lactate^11,12^ and the ketone body, β-hydroxybutyrate (BHB)^13^, both of which inhibit lipolysis through activation of closely related adipocyte-expressed G-protein coupled receptors (GPCRs), HCAR1 and HCAR2, respectively. The genes encoding these two receptors, as well as that for HCAR3, a very recently evolved family member which senses octanoate and is uniquely found in higher primates^14^, are closely co-located on a 30-kilobase stretch of chromosome 12q^15^. Variants in and around this locus have previously been reported to be associated with waist-hip ratio^5,16^ as well as circulating levels of triglycerides (TAG) and HDL cholesterol^17^ but it remains unclear which is the casual gene. We undertook studies to clarify the biological mechanism underlying these associations.

## Results

### The *HCAR* locus is associated with metabolic dysfunction

We first explored the locus using publicly available genome wide association study (GWAS) datasets from the Global Lipids Genetics Consortium (GLGC) and the Genetic Investigation of Anthropometric Traits (GIANT) consortium and noted robust associations of the *HCAR* locus with HDL cholesterol, triglycerides (TAG), and waist-hip ratio adjusted for BMI (WHRadjBMI) (Supplementary Figure 1). Further interrogation of the *HCAR* locus using UK Biobank (UKBB) exomes comprising ∼500K participants revealed that the strongest link was between *HCAR3 rs200905183 G>A* (p.R311C) and these metabolic traits (Supplementary Tables 1-3). However, we noted the high DNA sequence identity (97%) in the coding region between *HCAR2* and *HCAR3* (Supplementary Figure 2), which may pose challenges in accurately calling variants between the two genes using short read sequencing or genotype array data^18,19^, potentially explaining the substantial inconsistencies in the reported MAF for this variant between gnomAD and UKBB (Supplementary Table 4). We therefore developed a targeted genotyping approach to more accurately map the variant to *HCAR2* or *HCAR3*.

### Direct genotyping maps p.R311C to *HCAR2*

We designed a custom quantitative PCR-based genotyping assay with nested primers that could differentiate between *HCAR2* p.R311C (rs7314976 G>A) and *HCAR3* p.R311C (rs200905183 G>A, see methods and Supplementary Figure 3). We first genotyped 1,000 participants from the Fenland study^20^ for both possible variants and found a minor allele frequency for *HCAR2* p.R311C of ∼17%. In contrast, none of the 1,000 participants carried the putative *HCAR3* variant, suggesting that it is either non-existent or very rare, at least in European populations. We then proceeded to genotype 41.8K further individuals from Fenland and 3 additional studies - Ely^21^, EPIC-Norfolk^22^, and Oxford Biobank^23^ for the *HCAR2* p.R311C variant (Table 1).

**Table 1.**
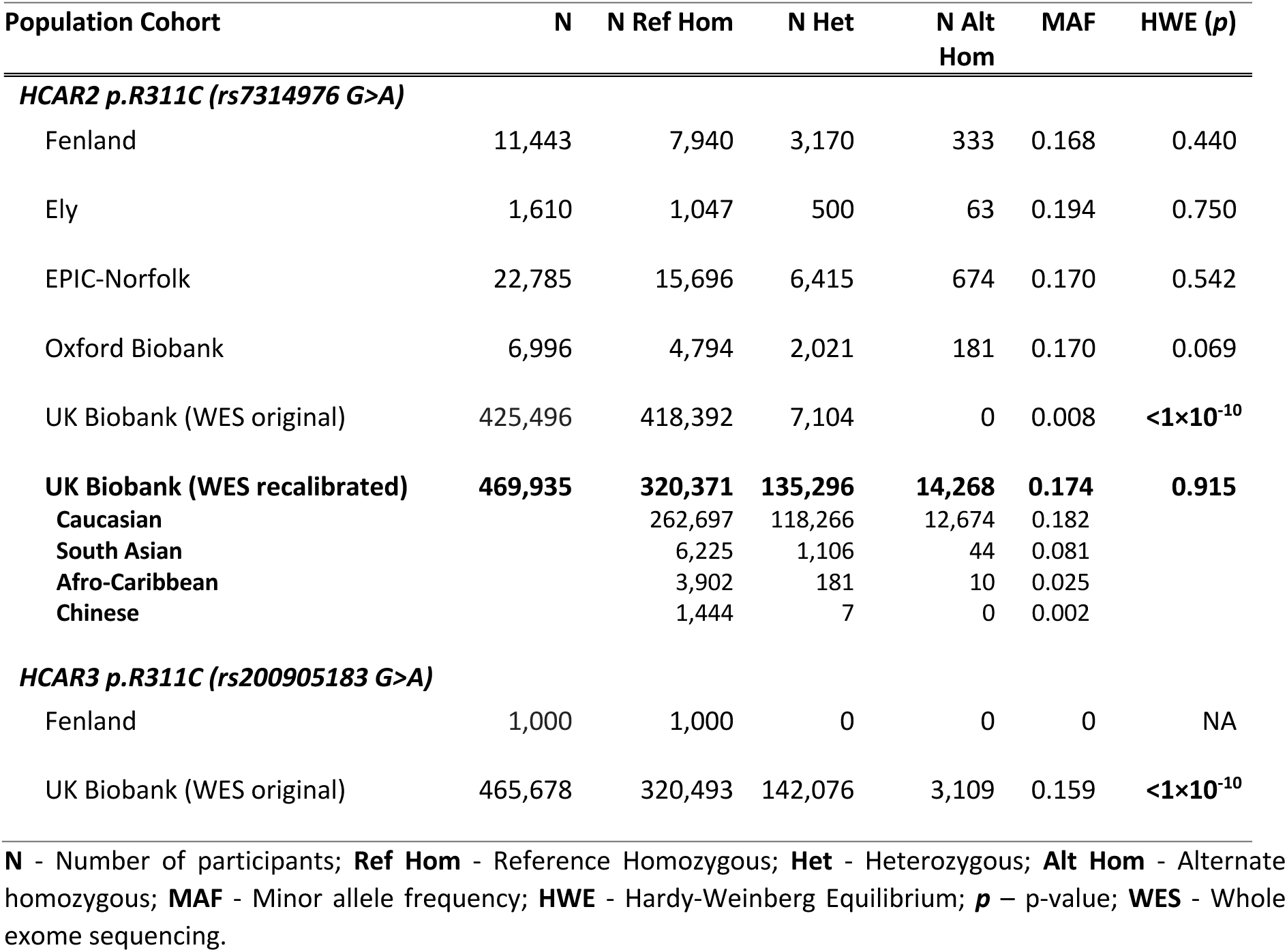
Genotype counts from UK population studies.

We also corrected the genotyping calls in the UKBB exomes by redistributing the reads with p.R311C alternate alleles from both genes to *HCAR2* and then remodelled the genotyping calls by minimising the HWE Chi-square (see methods, Supplementary Figure 4), listed in Table 1 (bold). This resulted in the observed MAF of 17.4% in the UKBB, which is comparable to the MAFs observed with custom genotyping in other cohorts, supporting the validity of this approach.

### HCAR2 p.R311C is associated with multiple adverse metabolic phenotypes in UK populations

We examined the association of the variant with anthropometric measures and selected metabolic phenotypes in the UKBB. Using the corrected genotype, we found that *HCAR2 p.R311C* was associated with increased WHRadjBMI (beta=0.046, 95%CI=0.040;0.051, p=1.5×10^-64^, Figure 1A, see Supplementary Table 5 for full results), consistent with the GWAS signal that originally highlighted the *HCAR* locus. This association was driven by both higher waist circumference (beta=0.012, 95%CI=0.009;0.014, p=3.10×10^-17^) and, more strongly, by lower hip circumference (beta=-0.017, 95%CI=-0.020;-0.014, p=1.80×10^-29^, Supplementary Table 5). *HCAR2 p.R311C* was also associated with lower BMI (beta=-0.136, 95%CI=-0.160;-0.112, p=1.4×10^-28^), which may mask some of the metabolically deleterious effects, and hence we used the BMI-adjusted model as the primary analysis.

**Figure 1:**
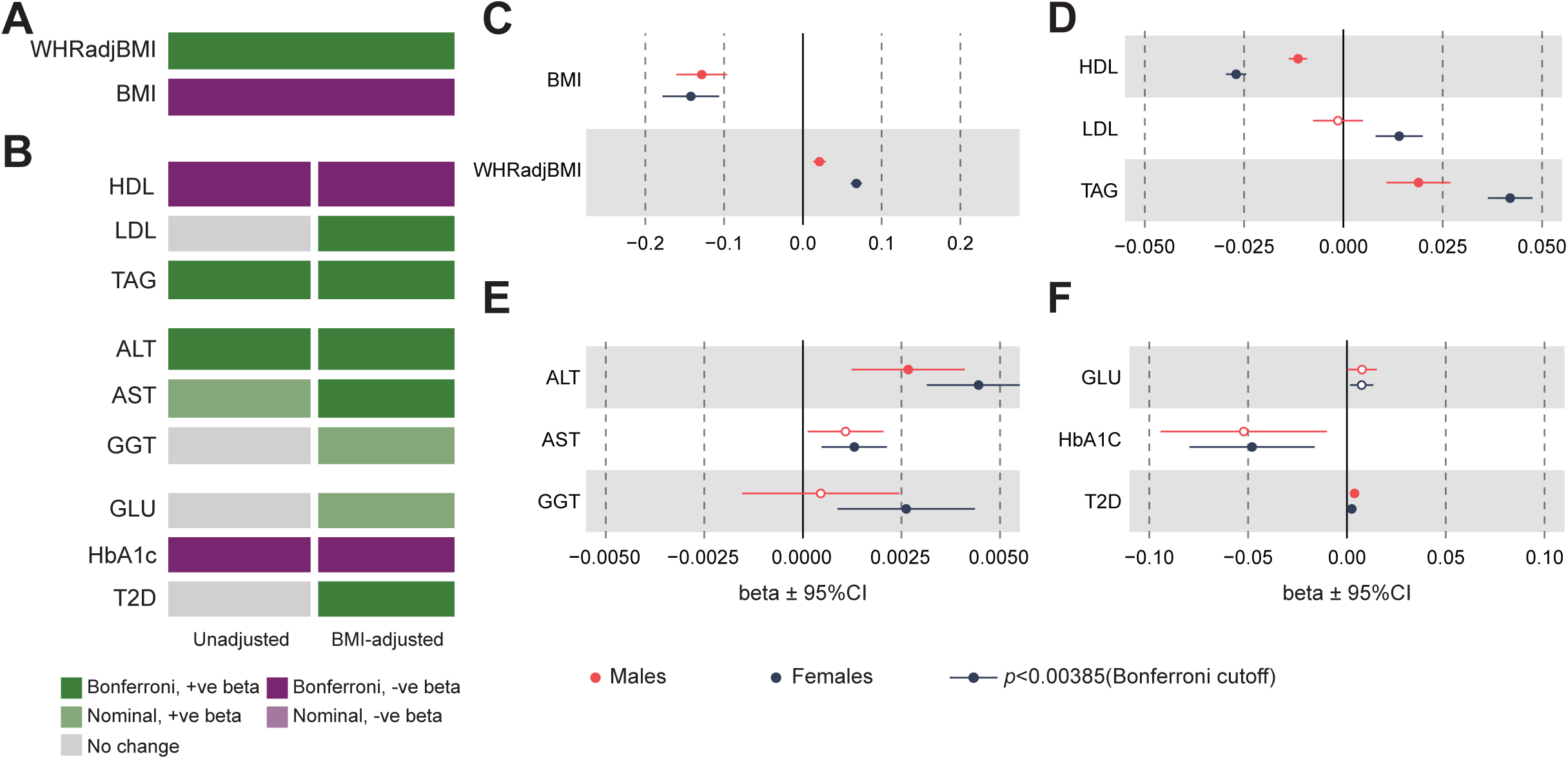
Genetic associations of *HCAR2 p.R311C* and metabolic traits in UK Biobank. (A-B) Heatmaps showing the association of *p.R311C* for (A) WHRadjBMI and BMI, (B) other metabolic phenotypes either unadjusted (left) and adjusted for BMI (right). (C-F) Sex-specific associations with anthropometric and metabolic phenotypes, all traits except BMI are adjusted for BMI.

*HCAR2 p.R311C* has adverse effects on lipid traits, with lower HDL cholesterol (beta=-0.019, 95%CI=-0.022;-0.018, p=2.10×10^-114^), higher TAG (beta=0.032, 95%CI=0.027;0.037, p=3.6×10^-39^), and a modest association with higher LDL (beta=0.0076, 95%CI=0.0033;0.0120, p=5.80×10^-4^) (Figure 1B). Variant carriers also had elevated liver enzymes: ALT (beta=0.0037, 95%CI=0.003;0.005, p=1.60×10^-13^), AST (beta=0.0013, 95%CI=0.0006;0.0019, p=8.70×10^-5^), and a nominal increase in GGT (beta=0.0016, 95%CI=0.0003;0.0029, p=1.60×10^-2^) (Figure 1B).

*HCAR2 p.R311C* was associated with increased T2D (OR=1.031, 95%CI=1.017;1.046, p=1.4×10^-5^) and higher random glucose levels (beta=0.007, 95%CI=0.002;0.012, p=3.90×10^-3^), but the effects are only seen in the BMI-adjusted model (Figure 1B). Interestingly, we also observed an unexpected association with lower HbA1c level (-0.055, 95%CI=-0.080;-0.029, p=2.90×10^-5^) (Figure 1B). While HbA1c correlates well with average plasma glucose levels, it could be influenced by red blood cell turnover.

In that regard, we noted a significant increase in reticulocyte count, a marker of increased red blood cell turnover, in carriers of the variant (beta=0.0007, 95%CI=0,0005;0.0010, p=7.7×10^-12^, Supplementary Table 5).

In sex-stratified analyses, *HCAR2 p.R311C* had a stronger effect on WHRadjBMI in females compared to males (Het p=4.37×10^-18^, Figure 1C). It also had stronger adverse effects on circulating lipid levels in females (HDL, LDL and TAG, Het p=6.7×10^-19^, 4.80×10^-4^ and 3.95×10^-6^, respectively, Figure 1D) but we observed no difference in the effects on circulating liver enzymes (Figure 1E). Effects on BMI (Figure 1C) and glycaemic traits (Figure 1F) were also similar between sexes (full results in Supplementary Table 5).

In a joint conditional analysis for the WHRadjBMI with *HCAR2 p.R311C* and 2 other common variant signals in proximity, namely *rs579877* (eQTL for *HCAR2*, see Supplementary Table 6); and *rs73226103* (∼10 kilobases downstream of *HCAR2 p.R311C*), we did not find any suggestion that the association of the recalibrated *HCAR2* p.R311C variant was attenuated after conditioning on the two other common variants (Supplementary Table 7).

We next performed a meta-analysis using directly genotyped data on ∼42K people from the Fenland, EPIC-Norfolk, Ely, and Oxford Biobank studies, where we have access to rich metabolic phenotypes. Consistent with the UKBB, *HCAR2 p.R311C was* associated with higher WHRadjBMI (beta=0.26, 95%CI =0.016;0.036, p=1.80×10^-7^, Figure 2B, association results in Supplementary Table 8) and lower BMI (beta=-0.029, 95%CI=-0.045;-0.014, p=2.10×10^-4^, Figure 2A). The WHRadjBMI effect was again driven by both decreased hip (beta=-0.022, 95%CI=-0.032;-0.012, p=1.02×10^-5^) and nominally increased waist circumference (beta=0.008, 95%CI=0.002;0.015, p=0.016) (Figure 2B), and BMI effect driven by modest reductions in both height and weight (Supplementary Table 8). The change in body fat composition was reflected in lower gynoid fat mass (beta=-0.030, 95%CI=-0.042;-0.018, p=4.09×10^-7^) and nominally higher android fat mass (beta=0.015, 95%CI 0.007;0.026, p=7.56×10^-3^) (Figure 2B). There was also evidence of decreased limb fat mass (beta=-0.038, 95%CI=-0.050;0.026, p=4.21×10^-10^) and nominally reduced arm fat mass (beta=-0.014, 95%CI=-0.025;-0.003, p=0.016) (Figure 2B).

**Figure 2:**
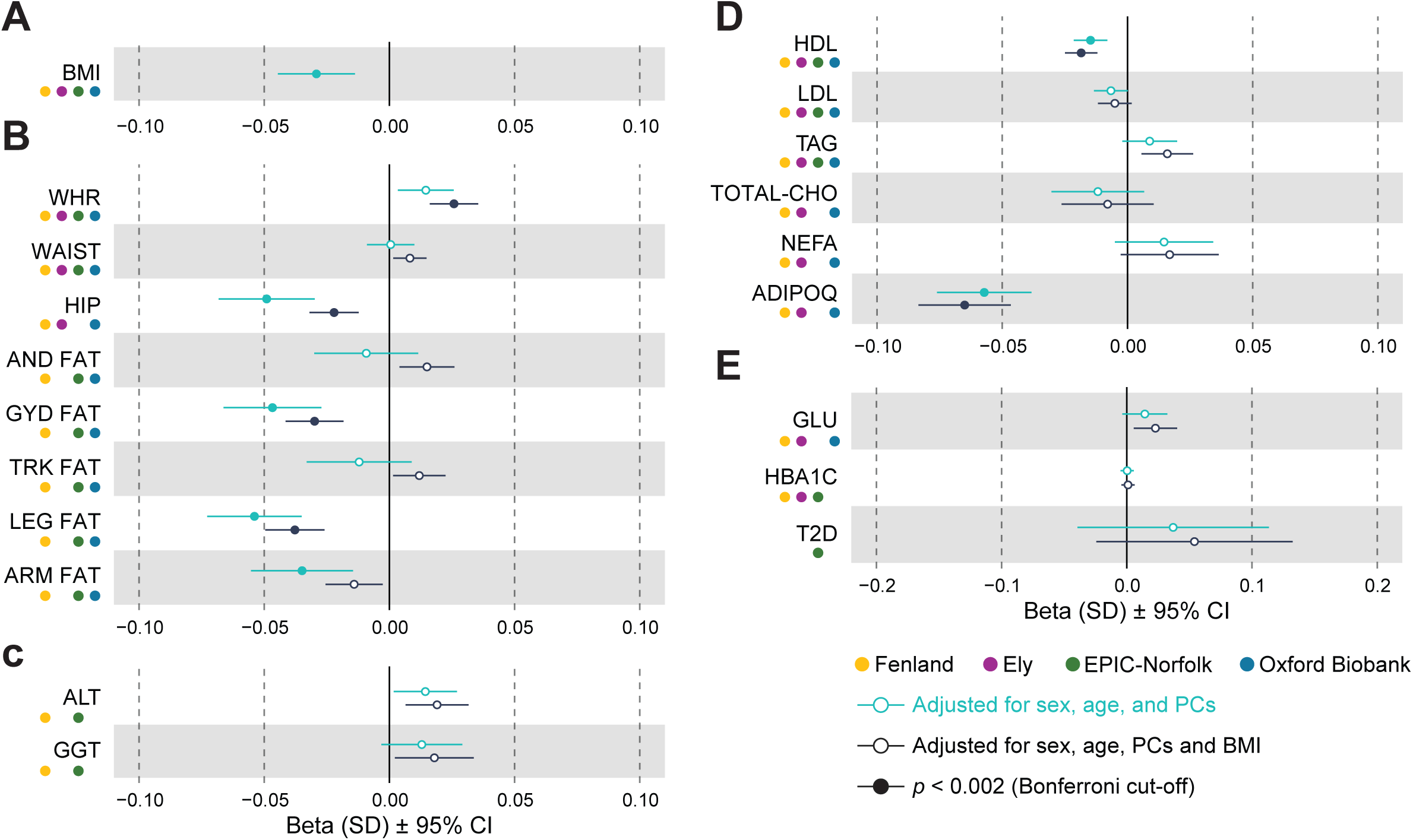
Meta-analysis of the association between HCAR2 p.R311C and metabolic traits in genotyped UK populations. In each case the figures show the results of a meta-analysis across the four studies with directly genotyped HCAR2 p.R311C adjusted for sex, age and first 10 principal components (turquoise) or additionally adjusted for BMI (black). (A) BMI and (B) Anthropometric phenotypes. WHR: Waist-to-Hip ratio; WAIST: Waist circumference; HIP: Hip circumference; AND FAT: Android fat mass; GYD FAT: Gynoid fat mass; TRK: Trunk fat mass; (C) Plasma lipid levels. (D) Blood glucose-related outcomes. GLU: Fasting Glucose; T2D: Type 2 diabetes. (E) Liver-related phenotypes. A Bonferroni cut-off = 2.0×10^-3^ was used for multiple testing correction.

Similar to UKBB, carriers also showed lower HDL cholesterol (beta=-0.018, 95%CI=0-0.025,-0.012, p=2.3×10^-8^), nominal increase in TAG (beta=0.016, 95%CI=0.006;0.026, p=0.0024), but we did not detect any effect on LDL (p=0.142) and total cholesterol levels (p=0.398) (Figure 2C). We also examined the effect of *HCAR2 p.R311C* on circulating non-esterified fatty acids (NEFA) levels, whilst there was no association with fasting NEFAs (p=0.091) (Figure 2C, we found a nominal increase in NEFA level at 120 minutes (p=0.022, n=827) post-oral glucose tolerance test (OGTT) in the Ely study, the only genotyped population in which this measure was available (Supplementary Figure 5, Supplementary Table 8).

*HCAR2 p.R311C* carriers had a lower circulating level of adiponectin (beta=-0.065, 95%CI=-0.083;-0.047, p=4.71×10^-12^), a circulating protein often used as a surrogate marker of insulin sensitivity^24^ (Figure 2C). In the cohorts in which data on plasma insulin both basally and 2 hour after oral glucose were available, *HCAR2* variant carriers had nominally higher levels of circulating insulin post-OGTT (beta=0.035, 95%CI=0.004;0.066, p=0.029, Supplementary Table 5) and increased basal and post-OGTT glucose levels (basal beta=0.023, 95%CI=0.006;0.040, p=0.009; post-OGTT beta=0.032, 95%CI=0.001;0,064, p=0.045), but there was no evidence of an effect on HbA1c levels (p=0.728), or incidence of type 2 diabetes (T2D, p=0.177) (Supplementary Table 8, Figure 2D).

The minor allele was nominally associated with elevated liver enzymes alanine transaminase (ALT) (beta=0.019, 95%CI=0.007;0.032, p=0.003) and gamma-glutamyl transferase (GGT, beta=0.018, 95%CI=0.002;0.037, p=0.026) (Figure 2E).

In summary, genetic association studies in five independent cohorts provided robust evidence that carriage of *HCAR2 pR311C* is associated with adverse alterations in fat distribution and markers of metabolic (glucose, lipid) and hepatic health. In the largest and most highly powered study (UKBB) it was also associated, after correction for BMI, with a significant increase in the risk of developing T2D.

### NMR Metabolome analysis of *HCAR2* p.R311C in the UK Biobank

We examined the association of *HCAR2* p.R311C with metabolites measured on the Nightingale platform in ∼280K participants of the UKBB. There were significant associations for 69 out of the 107 non-derived metabolites (Bonferroni cut-off = 0.000467, Supplementary Table 9) (Figure 3A). Consistent with the association with lower HDL cholesterol and higher TAG levels observed in the blood biochemistry data, *HCAR2* p.R311C was associated with reduction in multiple HDL associated metabolites and increase in several LDL and VLDL associated metabolites in the BMI-adjusted model (Figure 3A). Notably, concentrations of ketone bodies including acetone, acetoacetate and BHB were significantly lower in *HCAR2* p.R311C carriers (Figures 3A-B).

**Figure 3:**
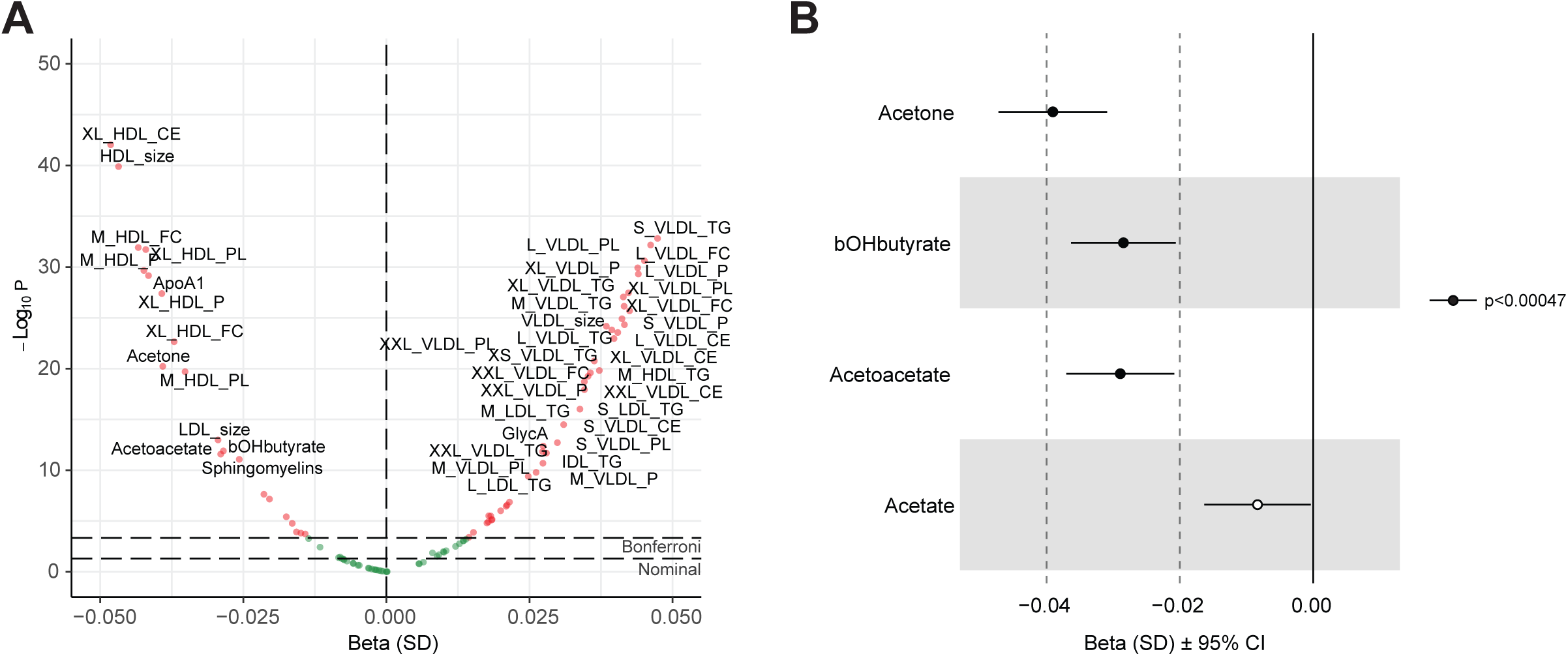
Effects of *HCAR2* p.R311C carriage on plasma metabolome in the UK Biobank. (A) Volcano plot showing differentially regulated plasma metabolites and (B) Effect estimates and 95% confidence intervals of circulating ketone bodies. Data are adjusted for sex, age and BMI.

### *HCAR2*, but not *HCAR3*, is expressed in adipose tissue

Having been concerned that the extremely close sequence homology between *HCAR2* and *HCAR3* might have confounded previous attempts to accurately determine sites of *HCAR2* gene expression in the human body, we designed gene-specific single-molecule in-situ hybridisation probes to differentiate clearly between *HCAR2* and *HCAR3* mRNA in various human tissues. HCAR2 expression in adipose tissue appeared heterogeneous, with high transcript abundance in a subset of adipocytes (Supplementary Figure 6A-B); expression was also detected in skin and seminal vesicles (Supplementary Figure 6B). HCAR3 was not detected in adipose tissue (Supplementary Figure 6B).

Of note, we did not find any expression of *HCAR2* or H*CAR3* in human pancreatic islets of Langerhans (Supplementary Figure 6B). This is consistent with the single-cell human islet atlas recently reported by Segerstolpe et al^25^.

### *HCAR2* p.R311C is enriched in patients with familial partial lipodystrophy

We hypothesised that *HCAR2* p.R311C might be associated with an increased risk of type 1 familial partial lipodystrophy (FPLD1)^26^. To test this, we obtained DNA from three different FPLD1 study cohorts from Spain, UK and France, and genotyped a total of 208 FPLD1 patients with 181 controls. We found that there was an overall over-representation of *HCAR2* p.R311C genotypes in the FPLD1 patients compared to control (OR=1.660, 95%CI=1.138;2.421, p=0.0085); however, this was driven exclusively by the enrichment in the Spanish cohort (Supplementary Table 10).

### Mice expressing *Hcar2* p. R308C have altered body composition and impaired metabolic function

To further explore the metabolic effects of HCAR2 p.R311C, we used CRISPR-Cas9 to generate a mouse model that expresses the homologous *Hcar2* p.R308C variant On chow diet, male mice homozygous for *Hcar2* p.R308C gained modestly more weight than WT littermate controls; this effect on body weight first appeared around 5-months of age and gradually increased before plateauing at 9 months of age (Figure 4A). While the body weight difference was non-significant at 5 months of age, assessment of body composition by EchoMRI revealed a significant increase in body fat mass in p.R308C mice (Figure 4B). Total lean body mass was comparable between WT and p.R308C mice (Figure 4C). The effect of the variant on body fat mass was further confirmed by MicroCT, which detected larger subcutaneous and visceral adipose tissue volume in the knock-in mice (Figure 4D-E).

**Figure 4:**
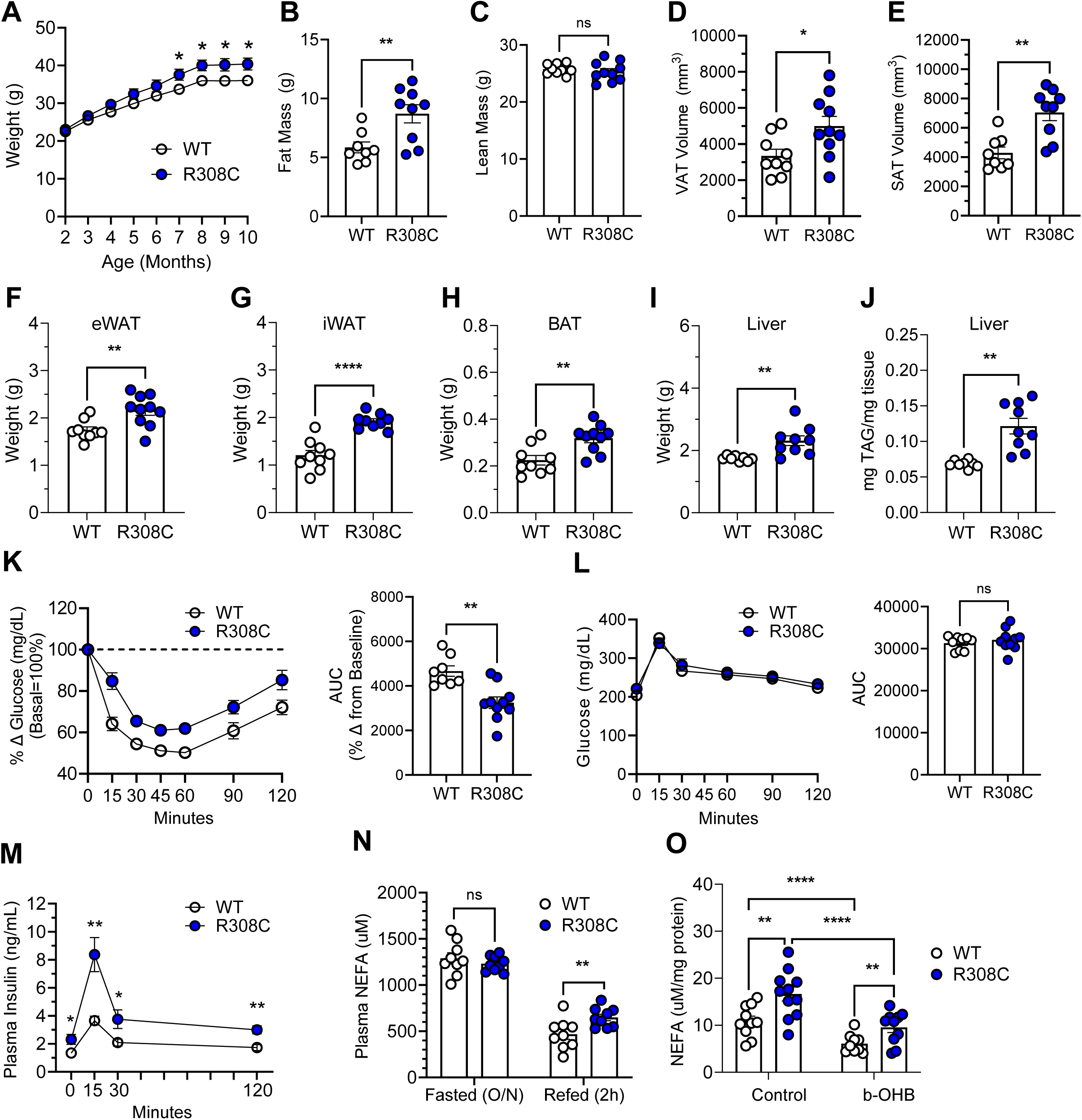
Metabolic phenotype of mice expressing the HCAR2 p.R308C variant. (A) Body weight gain of mice expressing HCAR2-WT (WT, hollow) or HCAR2-R308C (R308C, blue) on chow diet from 2 months to 10 months of age; unpaired t-test used to compare genotypes at each timepoint. (B) Fat mass, and (C) lean mass of WT and R308C mice at 5 months of age measured by EchoMRI; unpaired t-test. (D) Visceral, and (E) Subcutaneous adipose tissue volume of WT and R308C mice at 6 months of age measured by MicroCT; unpaired t-test. (F) Epididymal adipose tissue weight, (G) inguinal adipose tissue weight, (H) brown adipose tissue weight, and (I) liver weight of WT and R308C mice at 12 months of age; unpaired t-test. (J) Liver triglyceride content of WT and R308C mice; Welch’s t-test. (K) Left: Insulin tolerance of WT and R308C mice at 9 months of age. Data is presented as a percentage of basal glucose. Right: Quantification of area under the curve, unpaired t-test. (L) Left: Oral glucose tolerance of WT and R308C mice at 9 months of age. Right: Quantification of area under the curve; unpaired t-test. (M) Glucose-stimulated insulin secretion of WT and R308C mice; Welch’s t-test used to compare genotypes at each timepoint. (N) Plasma NEFA levels of WT and R308C mice in the fasted state (overnight, O/N), and the re-fed state (2-hours post feeding); 2-way ANOVA with multiple comparisons test. (O) NEFA concentration per milligram of eWAT isolated from WT and R308C mice; data is shown for both basal activity and activity following acute stimulation with 10 mM BHB. For A-N, N=9 WT and 10 R308C males. Body weight and EchoMRI findings have been in repeated in an additional cohort. For ex vivo lipolysis in O, experiment has been performed 2 times using one fat pad per condition. All values presented as mean ± SEM. *P < 0.05, **P < 0.01, and ***P < 0.001.

The effect of p.R308C on body composition was maintained at 9 months of age (Supplementary Figure 7A-B), and, at necropsy, the p.R308C mice had significantly heavier eWAT, iWAT, BAT, and liver tissue weights (Figure 4F-I). Although there was no discernible difference in plasma cholesterol or TAG (Supplementary Figure 7C-D) between groups, the p.R308C mice had a significant (∼2-fold) increase in liver TAG (Figure 4J), accompanied by a trending increase in plasma ALT and AST (Supplementary Figure 7E-F). The effect of *Hcar2* p.R308C on body weight, tissue weights, and liver TAG are all consistent with published findings in HCAR2-knockout mice, providing evidence that p.R308C is likely to be a loss-of-function mutation in vivo^27,28^.

The increased fat mass of p.R308C mice was accompanied by a significant defect in the insulin response relative to littermate controls (Figure 4K). The p.R308C mice had no defect in oral glucose tolerance (Figure 4L); however, p.R308C mice had increased fasting insulin and secreted significantly more insulin following oral administration of glucose, likely a compensatory response for insulin resistance (Figure 4M).

We next investigated the regulation of lipolysis by WT and p.R308C mice. While there were no differences in fasting NEFA levels, consistent with what is seen post-glucose load in humans carrying the variant, mice expressing *Hcar2* p.R308C have higher NEFA levels two hours after refeeding (Figure 4N). Lastly, since an increase in basal lipolysis is associated with impaired glucose metabolism^29^, we examined basal lipolytic activity in adipose tissue explants from WT and p.R308C mice. NEFA levels in the medium of p.R308C explants were significantly higher than that of control tissue under unstimulated conditions, indicative of increased basal lipolytic activity. Following treatment with BHB, WT and p.R308C explants showed a similar degree of lipolytic suppression; however, the NEFA levels still remained significantly higher in the media of variant explants (Figure 4O).

### Pharmacological profiling of HCAR2 p.R311C reveals enhanced basal signalling activity and reduced receptor surface availability

To study the signalling properties of the *HCAR2* p.R311C variant, we utilized a BRET-based biosensor platform (bioSens-All®, Domain Therapeutics), which enables quantification of the GPCR signalling response at multiple points in the pathway. Using this approach, HEK293 cells transiently transfected with either *HCAR2*-WT or *HCAR2*-<u>p.</u>R311C were stimulated with one of six HCAR2 agonists (nicotinic acid (NA), BHB, GSK-256073, MK-1903, MK-0354, and monomethyl fumarate (MMF)). Because HCAR2 is a Gi-coupled receptor, we first investigated whether the p.R311C variant has altered Gi1 coupling activity. Interestingly, the p.R311C variant exhibited a modest gain-of-function in basal Gi1 activity (Emin). The Emin was consistently elevated across all experimental settings, while a significant increase in maximal activity (Emax) was found with three compounds (NA, GSK-256073, and MK-0354). (Figure 5A-D and Supplementary Figure 8A-B; quantification and statistics in Supplementary Table 11). We observed a trending decrease in EC50 for p.311C, which only reached significance for MK-1903, suggesting the increase in activity is driven largely by constitutive basal activity.

**Figure 5:**
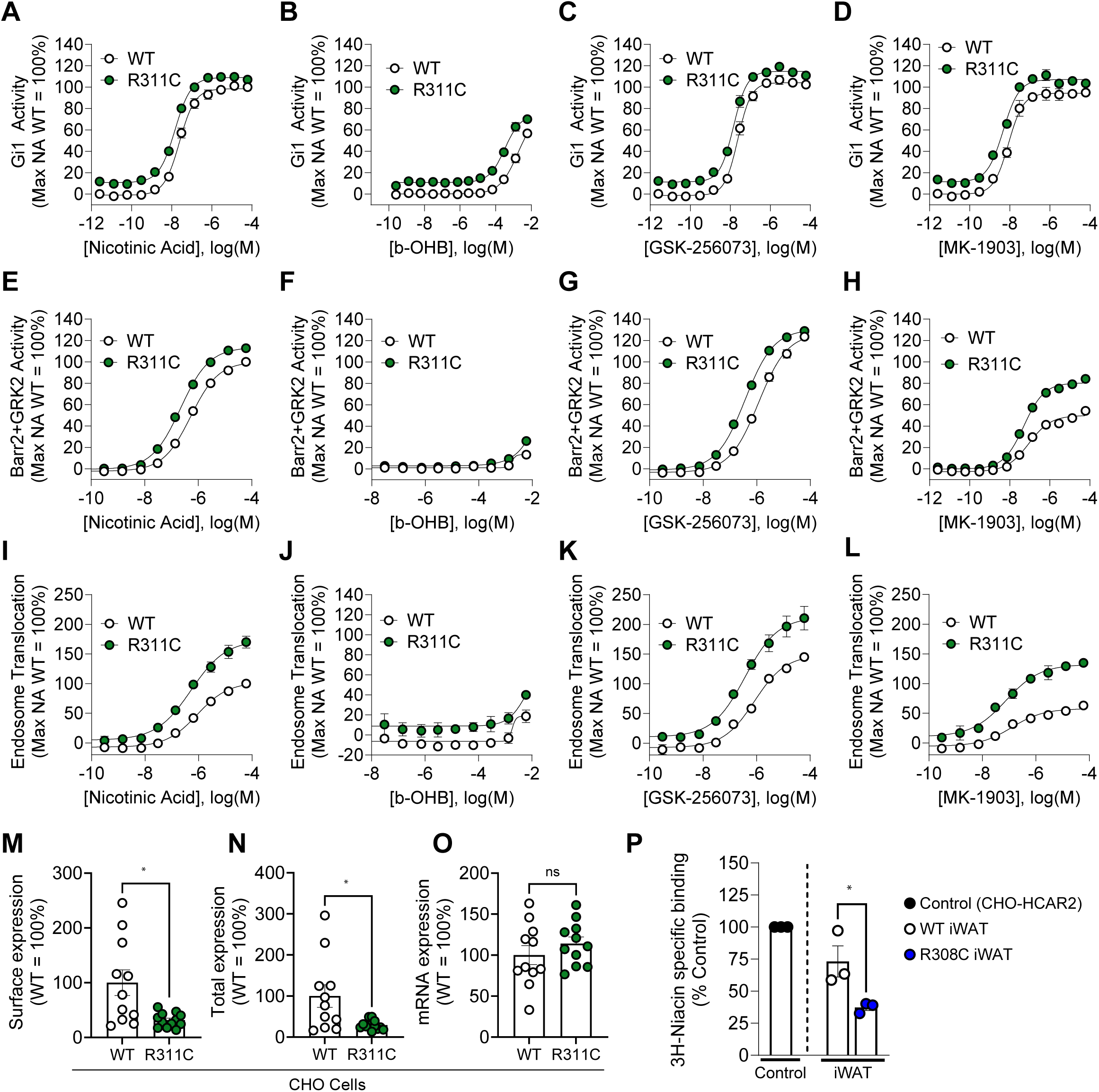
Molecular characterisation of HCAR2 p.R311C receptor activity and localization. (A-D) Gi1 signalling activity of HEK 293 cells transiently transfected with *HCAR2-*WT (hollow) or *HCAR2-*p.R311C (dark green). Cells were stimulated with (A) nicotinic acid, (B) BHB, (C) GSK-256073, or (D) MK-1903 for 10 minutes prior to measurement of activity using a BRET-based biosensor. Data is normalized to the percent maximum activity of nicotinic acid. EC50, E_min_, and E_max_ with corresponding statistics can be found in Supplementary Table 11. (E-H) β-arrestin 2 and GRK2 activity of HEK 293 cells transiently transfected with *HCAR2-*WT or *HCAR2-*p.R311C. Cells were stimulated with (E) nicotinic acid, (F) BHB, (G) GSK-256073, or (H) MK-1903 for 10 minutes prior to measurement of activity using a BRET-based biosensor. Data is normalized to the percent maximum activity of nicotinic acid EC50, E_min_, and E_max_ with corresponding statistics can be found in Supplementary Table 12. (I-L) Early endosome translocation activity of HEK 293 cells transiently transfected with *HCAR2-* WT or *HCAR2-*p.R311C. Cells were stimulated with (I) nicotinic acid, (J) BHB, (K) GSK-256073, or (L) MK-1903 for 30 minutes prior to measurement of activity using a BRET-based biosensor. Data is normalized to the percent maximum activity of nicotinic acid EC50, E_min_, and E_max_ with corresponding statistics can be found in Supplementary Table 13. (M) Surface expression of HCAR2 in CHO clonal cell lines stably expressing Hibit-HCAR2-WT or Hibit-HCAR2-p.R311C. Data is presented as percent of WT Maximum and is shown for 12 individual clones; Welch’s t-test. (N) Total (whole cell) expression of HCAR2 in cells from (M); Welch’s t-test. (O) mRNA expression of *HCAR2* of cells from (M); unpaired t-test. (P) Specific binding of [3H]-Niacin to membranes isolated from iWAT fat pads of mice expressing *Hcar2-*WT or *Hcar2*-p.R308C. Data is normalized to a representative CHO clonal cell line expressing *HCAR2-*WT (Control); unpaired t-test used to compare binding to WT (hollow) and p.R308C (blue) membranes. For A-L, all experiments have been performed 3 separate times, and the average of 3 experiments is shown. M-O have been performed 4 separate times, and the average of 4 experiments is shown. P has been performed 3 separate times, and the average of 3 experiments is shown. Similar results have also been achieved from mouse visceral adipose tissue. All values presented as mean ± SEM. *P < 0.05.

We also observed an enhanced recruitment of β-arrestin 2 and GRK2 with the p.R311C variant, similar in magnitude to the increase in Gi1-coupling (Figure 5E-H and Supplementary Figure 8C-D; quantification and statistics in Supplementary Table 12), indicative of increased susceptibility to desensitization of the p.R311C variant. Supporting this notion, we found that the p.R311C variant had a robust increase (134.9% at 0.65 uM NA) in the recruitment to early endosomes after a 30-minute agonist stimulation (Figure 5I-L and Supplementary Figure 8E-F; quantification and statistics in Supplementary Table 13). In summary, the BRET-based biosensor data suggested that enhanced basal activity of the p.R311C variant may lead to premature desensitization and internalization of the receptor.

We next evaluated if the enhanced desensitization and internalization of the p.R311C variant had subsequent consequences on receptor expression. To this end, we generated CHO clonal cell lines that stably express either Hibit-tagged *HCAR2*-WT or *HCAR2*-p.R311C constructs. We found that clonal lines harbouring the p.R311C variant consistently expressed lower levels of receptor both at the cell surface and the whole cell level (Figure 5M-N). These findings were consistent across twelve different clones each for WT and p.R311C. A similar decrease in expression was observed by immunoblotting using whole cell lysates (Supplementary Figure 8G). Notably, reduced steady-state levels of HCAR2-R311C protein were not accompanied by any change in the levels of *HCAR2* mRNA, implying a post-transcriptional effect (Figure 5O). To evaluate whether this finding translated *in vivo,* we isolated cell membranes from iWAT of WT and p.R308C mice and used radiolabelled niacin to perform a specific binding assay (Figure 5P). Membranes from a CHO clonal cell line expressing Hibit-HCAR2 WT was used as a positive control. Consistent with the *in vitro* results, we measured a ∼50% reduction in 3H-niacin binding to the p.R308C membranes, indicative of reduced receptor availability at the cell surface.

## Discussion

Ketone bodies play an important role in normal physiology, providing a source of energy, particularly to the brain, during periods of fasting, prolonged exercise, or when carbohydrate intake is low^30^. Diets that deliberately provoke ketogenesis are used therapeutically in certain neurological conditions^31^ and have been extensively evaluated for their impacts on metabolic health^32^. Dietary ketone supplements are in widespread use, though the extent to which they are beneficial is not yet securely established^33^. In addition to being metabolised intracellularly, at least one of the main ketone bodies, β-hydroxybutyrate (BHB), signals via the cell surface-expressed G protein coupled receptor (GPCR), HCAR2. In adipocytes, activation of HCAR2 leads to an inhibition of lipolysis, which serves to exert negative feedback control on ketone production, as free fatty acids released from adipocytes are the major substrate for ketone production by the liver. This likely contributes to the fact that while circulating ketone body and NEFA levels rise steeply with starvation, steady state levels of both are reached before dangerously acidotic levels of ketone bodies are reached^34^.

Our studies have established that a common missense variant in the C-terminal tail of HCAR2 chronically impairs HCAR2 signalling and is associated with features of the metabolic syndrome in humans and in mice. Although it is formally possible that HCAR2 expression in other tissues contributes to this phenotype, it is most likely that a defect in the adipocyte control of lipolysis is key to the dysmetabolic phenotype seen in vivo.

The unusually extensive DNA sequence identity between *HCAR2* and its recently evolved and neighbouring paralog, *HCAR3*, hindered the efforts in identifying the casual gene within this *HCAR* locus. The development of a targeted genotyping assay was hence required to unequivocally determine the attribution of the R311C exclusively to *HCAR2*. While it remains possible that carriers of an *p.R311C* variant in *HCAR3* exist in some human populations, we did not find it in 1000 white Europeans, a population in which the prevalence of the variant in *HCAR2* was 17%.

Knowledge of the correct location of *p.R311C* allowed us to re-analyse the association of the variant with anthropometric and metabolic variables at scale in UKBB and, in parallel, examine relevant associations in four directly genotyped UK cohorts, which, although considerably smaller, provided the ability to analyse additional relevant phenotypes. Across all cohorts, carriage of *p.R311C* is robustly associated with increased waist hip ratio (driven primarily by a reduction in hip circumference), elevated triglycerides and lower circulating concentrations of HDL. A somewhat more variable association is seen with elevated circulating levels of liver enzymes. Although there was no significant association with fasting levels of NEFAs, in the one study in which NEFAs were measured after a glucose load, carriage of the variant was associated with impaired suppression of NEFAs, consistent with what was found in mice carrying the homologous variant. The variant is associated with a small decrease in BMI and, after adjusting for that, the *p.R311C* variant is significantly associated with the risk of T2D in the ∼500K participants of UKBB. While we cannot entirely exclude a collider effect, the broad associations with adverse fat distribution and circulating lipids in both BMI-adjusted and unadjusted models strongly suggest that HCAR2 likely plays a role in maintaining metabolic health.

Impaired suppression of adipocyte lipolysis might be expected to result in increased ectopic storage of fat, resulting in insulin resistance. Mice homozygous for murine equivalent of HCAR2 R311C were insulin resistant on direct testing and, in human carriers, serum adiponectin, often used as a surrogate for insulin resistance, was significantly reduced. Unfortunately, in UKBB, there is no direct measure of insulin resistance, so our study of insulin-related phenotypes was limited in size and power. However, in the cohort studies where fasting and post prandial insulin measurement were available, levels were somewhat higher in the variant carriers.

Despite the association of the variant with measures of metabolic dysfunction including higher glucose levels, HbA1c levels in UKBB participants were actually significantly lower in variant carriers. HbA1c is influenced not only by ambient glucose levels but also by red blood cell turnover rate. We noted that the carriers of p.R311C in UKBB also had significantly elevated reticulocyte percentage and reticulocyte counts, indicating a higher rate of red blood cell turnover. A recent study has identified an additional possible role for HCAR2 as a cell surface receptor for heme that is upregulated during states of increased haemolytic activity^35^. The involvement of HCAR2 in this aspect of haematological regulation may explain, at least in part, both the increased reticulocyte production and the paradoxically lower HbA1c seen in carriers of p.R311C.

The finding of lower levels of circulating ketone bodies in variant carriers in UKBB may seem surprising. In many situations where an endocrine GPCR is defective, (for example the TSH receptor) circulating levels of its endogenous ligand are elevated. While more research on the production and clearance of ketone bodies in human and murine carriers will be needed to provide a definitive explanation for the lower levels, we speculate that a state of insulin resistance/hyperinsulinemia that carriers are prone to develop would be likely to activate hepatic lipogenesis at the expense of fatty acid acid oxidation^36^. Thus, even though the excessive adipocyte lipolysis would produce more potential substrate of hepatic ketogenesis, the hepatocyte pathways for the synthesis of ketones, which are products of fatty acid oxidation, may be partially inhibited. This is consistent with previous studies which have reported lower circulating ketone body levels despite higher circulating NEFA levels in individuals with obesity^37^.

In order to provide orthogonal testing of the impact of the variant on *in vivo* metabolism we generated and characterised a mouse engineered to express the homologous mutation to p.R311C in *Hcar2*. As mice are generally more metabolically tolerant of primary perturbations in adipocyte function than humans^38^, we chose to study mice that were homozygous for the mutation. Consistent with what we found in humans, mice carrying the variant showed impaired suppression of post-prandial NEFA levels and adipose explants from the mice showed increased NEFA release, both basally and after exposure to BHB. The mice were hyperinsulinemic after a glucose load and resistant to exogenous insulin. Previous reports of mice lacking any functional *Hcar2* describe an increase in fat mass and liver TAG^27,28^, a phenotype which we also observed in our mice, which provides further evidence that the R311C mutation results in reduced receptor signalling. In contrast, human carriers of p.R311C have a significantly reduced BMI and total fat mass which implies some cross-species differences in the role of HCAR2 in the control of energy balance.

Given that both the human genetics and mouse model phenotype suggested *HCAR2* p.311C is a loss-of-function variant, we were initially surprised that the BRET-based biosensor results revealed an acute gain-of-function in Gi-signalling for the variant. However, this gain-of-function is largely driven by a significant increase in the Emin, and not a difference in the EC50, suggesting the variant has high basal/constitutive activity. Furthermore, the compensatory increase in both β-arrestin recruitment and early endosome translocation provide evidence that this gain-of-function is likely transient. This is supported by our findings that the mutant receptor expression is reduced at the cell surface both *in vitro* and in mouse adipose tissue. It is likely that chronic low-level activation of HCAR2 by endogenous BHB and/or dietary niacin drives continuous receptor internalization, resulting in a net loss-of-function *in vivo*. A similar phenomenon has been observed for other GPCRs implicated in metabolic health; for example, the common glucose-dependent insulinotropic polypeptide receptor (GIPR) E354Q variant has enhanced basal activity and an agonist-induced internalization rate 2.1-2.3 times faster than WT GIPR. The E354Q variant’s altered kinetics reduce receptor expression at the surface and impair the response to GIP, explaining the increased risk of Type 2 diabetes among carriers^39,40^.

Consistent with the reduced surface expression of p.R311C, a previous study demonstrated that deletion of HCAR2 residues 295-314 impairs receptor localization to the cell surface^41^. Although the mechanism by which HCAR2 p.R311C drives receptor desensitization and internalization is unclear, the unaltered levels of *HCAR2* mRNA indicate that the effect is occurring post-translationally. One possibility is the alteration of post-translational modifications, such as palmitoylation. Cysteine residues are a common site for palmitoylation, a modification known to alter receptor trafficking amongst cellular compartments^42–44^ at the cell membrane. Future molecular studies of this domain and the specific impact of this variant on the receptor structure, should provide useful additional insights for modelling the effect of this common mutation. HCAR2 has previously garnered interest for the pharmacological treatment of dyslipidaemia. In 2003, it was discovered that niacin exerts its beneficial effects on circulating TAG and HDL through adipocyte HCAR2^45–47^, prompting the development of synthetic HCAR2 agonists (MK-1903, MK-0354, and GSK-256073) that harness the beneficial effects of niacin without inducing cutaneous flushing. However, clinical development of these agonists was discontinued due to lack of sustained efficacy in patients^48,49^. Whether the tachyphylactic effect was more pronounced – or partially driven by – HCAR2 p.R311C carriers warrants further investigation.

The effect of R311C on receptor function is relatively subtle which likely explains why this variant has reached such a high frequency in the population. We initially found the R311C variant in UK European populations, where it appears to have a MAF of ∼17%. Using corrected genotyping data from UKBB we found a somewhat lower prevalence (∼8%) in participants of south Asian origin. The variant appears to be rare in individuals of African or African Caribbean descent (Table 1). The extent to which more damaging variants, including those found in homozygosity, contribute more broadly to human metabolic dysfunction remains to be explored. Such studies will need to address the technical challenge of discriminating between variants in the *HCAR2* and *HCAR3* genes.

In summary, we have found that a variant in the receptor for β-hydroxybutyrate, which impairs ketone body signalling, is common in some major human populations. Importantly, it is associated with impaired control of lipolysis in adipose tissue with the resulting likely excess of ectopic fat predisposing to dyslipidaemia, hepatic fat accumulation, insulin resistance and Type 2 diabetes. These data support the idea that ketone body sensing by adipocytes plays an important role in the maintenance of human metabolic health.

## Methods

### Population study cohorts

#### UK Biobank

The UKBB is a large and prospective study of approximately 500,000 healthy participants aged 40–69 years, recruited between 2006 and 2010^50^. All analyses conducted using the UKBB Resource were done under application numbers 9905, 32974, and 26041.

Phenotypes examined in this article include BMI, WHRadjBMI, HDL and LDL cholesterol, triglycerides, ALT, AST and GGT, glucose, HbA1c, reticulocyte and erythrocyte counts, T2D defined as in Gardner et. al ^51^ and serum metabolomic measures (non-derived only) as measured by the Nightingale NMR metabolomic profiling platform. Detailed information for phenotypes could be found at the UK Biobank Showcase website (https://biobank.ndph.ox.ac.uk/showcase/).

#### The Fenland study

The Fenland study is a population-based cohort of 12,435 participants born between 1950 and 1975 who underwent detailed phenotyping at the baseline visit between 2005 and 2015. The study was approved by the Cambridge Local Research Ethics Committee and all participants provided written informed consent. In brief, the participants were recruited from general practice surgeries in the Cambridgeshire region in the UK. Individuals were not enrolled in the cohort if they were clinically diagnosed with diabetes mellitus, a terminal illness or psychotic disorder, were unable to walk unaided, or were pregnant or lactating.

Phenotypes used in this article: anthropometric variables including BMI, waist and hip circumferences, WHR, and fat depot measurements from dual-energy X-ray absorptiometry (DEXA); Blood biochemistry including total, HDL and LDL cholesterol, triglycerides, adiponectin, HbA1c, ALT and GGT, fasted and 120-min post-OGTT non-esterified fatty acids (NEFA), glucose and insulin. Please refer to the study data dictionary (https://epidata-ext.mrc-epid.cam.ac.uk/ddic/overview/Fenland/) for further information.

#### The EPIC-Norfolk study

The EPIC-Norfolk study is a prospective cohort of 25,639 healthy individuals aged between 40 and 79 years and living in the county of Norfolk in the UK at recruitment. The study was approved by the Norfolk Research Ethics Committee and all participants gave their written consent before entering the study.

Phenotypes used in this article: BMI, waist circumference, imputed DEXA measurements derived from bioelectrical impedance^52^, blood biochemistry including HDL and LDL cholesterol, triglycerides, ALT, GGT and T2D. Please refer to the study data dictionary for further information (https://www.epic-norfolk.org.uk/cgi-bin/dictionary?FMT=topterms).

#### The Ely study

The Ely study is a longitudinal study established in 1990 to study the aetiology and incidence of diabetes and metabolic disorders among individuals without clinically diagnosed diabetes at baseline. Around 1,000 Individuals free from diabetes in 1990 were studied at 5 and 10 years with detailed measurement of glucose tolerance and metabolic characterisation with particular emphasis on the quantification of energy expenditure. The study was approved by the local research ethics committee, and all participants gave written informed consent.

Phenotypes used in this article: BMI, waist and hip circumferences, WHR, blood biochemistry including total, HDL and LDL cholesterol, adiponectin, fasted and 120-minute post-OGTT plasma NEFA, glucose and insulin, and HbA1c. Further information could be found from the study data dictionary (https://epidata-ext.mrc-epid.cam.ac.uk/ddic/overview/Ely/)

#### Oxford Biobank

The Oxford Biobank was healthy, recallable population cohort of 30-50 years old in Oxfordshire UK for genomics translational research with a focus on metabolic diseases. The study has collected a broad range of metabolic-, CVD- and obesity-related phenotypes based on blood plasma phenotyping, genetic biomarkers, questionnaires, anthropometric measurements and body composition assessment using DEXA. The study received ethical approval from Oxfordshire Clinical Research Ethics Committee, and all subjects gave written informed consent. Further information on the measurements could be found in the cohort profile article^23^.

#### Spanish FPLD1 cohort

This study was approved by the ethics review panel of the Red Gallega de Comités de Ética de la Investigación (approval code 2017/477).

### SNP Genotyping of HCAR2 rs7314976 G>A and HCAR3 rs200905183 G>A by Quantitative PCR

Genotyping of rs7314976 and rs200905183 was performed using custom rhAmp™ SNP genotyping assay (Integrated DNA technologies), design number CG.GT.XFZV8105.1 and Hs.GT.rs200905183.A.1 respectively. Each assay consists of 3 primers: ASP1 for wildtype allele, ASP2 for alternative allele and a locus specific primer (LSP), the detail design is depicted in Supplementary Figure 3. Quantitative PCR was performed on a Thermo Quantstudio 5 instrument (Thermofisher) with the following cycling condition: 95°C for 10 min, 40 cycles of 95°C for 10 seconds,60°C for 30 seconds, 68 °C for 20 seconds and a final step of 68 °C for 30 seconds. The allele detection was via Fam for ASP1 (rhAmp-F) and Yakima Yellow for ASP2 (rhAmp-Y).

### Linear regression analyses in Fenland, EPIC-Norfolk, Ely and Oxford Biobank

All quantitative phenotypes were standardised to a mean of 0 and standard deviation of 1. The association analysis for Fenland, EPIC-Norfolk, and Ely was performed using lm() function in R (4.3), with sex, age, first 10 principal components, with or without BMI included as covariates. T2D variable in EPIC-Norfolk was derived as described by Cardona et al.^53^. The regression analysis in Oxford biobank was run using PLINK version 1.90b3e, adjusted for age, sex, first 4 principal components, plus with or without adjustment for BMI. Bonferroni correction was used to account for multiple testing.

### Meta Analysis

Meta analysis for genotyped studies was performed using meta (8.0-1) package in R (4.4) using the summary statistics (beta and standard errors) from each individual study. Bonferroni correction was used to account for multiple testing.

### Recalibration of HCAR2 rs7314976 G>A calls in UK Biobank

For each participant within the UK Biobank whole-exome sequence (470K) dataset, the reference and alternative allele read counts from both *rs7314976* and *rs200905183* were first aggregated and then merged into a new combined site (*HCAR2/3* p.R311C). Given that rs200905183 was extremely rare (<1:1000) in the Fenland study, we assumed that all the alternative allele sequence reads would have originated from *rs7314976* (*HCAR2*) and the equivalent site in *HCAR3* would only contain the reference allele reads. Based on this assumption, homozygous and heterozygous genotypes at rs7314976 G>A are expected to have allele balance (AB) of around 50% and 25%, respectively.

For the recalibration of genotyping calls for rs7314976 G>A, we performed a stepwise modelling of genotypes (0/0, 0/1, 1/1) by raising AB cutoff for calling heterozygous (0/1) and homozygous (1/1) genotypes from 0.2 to 0.5, with increments of 0.001 at each step. Next, we fitted the genotyping counts 0/0, 0/1 and 1/1 from all individuals in the dataset into the Hardy-Weinberg Equilibrium (HWE) and calculated the chi-square. The AB that resulted in the lowest HWE chi-square (AB=0.41) was chosen as the final cutoff.

### Phenotype Association Analysis in the UK Biobank

All the variables were quality checked for normality and extreme values (≥2SD) were removed. Liver enzymes ALT, AST and GGT were log2-transformed before the analysis. The association testing in the UK Biobank was performed using bolt-lmm (2.3.6 and 2.4.1) infinitesimal model, which allows the inclusion of related individuals, with sex, age, first 10 principal components provided by the UKBB, with and without BMI as covariates.

### Conditional analysis

The conditional analysis for HCAR2 variants was performed using Genome-wide Complex Trait Analysis (GCTA)-COJO (version 1.92.0 beta2) and a window size of 501Kb to identify independent associations of individual SNPs.

### Single-molecule in-situ hybridisation

Detection of *HCAR2/HCAR3* targets as well as positive and negative control targets was performed on FFPE sections using Advanced Cell Diagnostics (ACD) BaseScope LS Reagent Kit (Cat. No: 323600) and various BaseScope LS probes (see table below). (ACD, Hayward, CA, USA). Briefly, sections were cut at 5 microns, baked for 1 hour at 60°C before loading onto a Bond RX instrument (Leica Biosystems). Slides were deparaffinized and rehydrated on board before pre-treatments using Epitope Retrieval Solution 2 (Cat No. AR9640, Leica Biosystems) at 88°C for 15 minutes, and ACD Enzyme from the LS Reagent kit at 40°C for 10 minutes. Probe hybridisation and signal amplification was performed according to manufacturer’s instructions. Fast red detection of each target was performed on the Bond Rx using the Bond Polymer Refine Red Detection Kit (Leica Biosystems, Cat No. DS9390) according to ACD protocol. Slides were then removed from the Bond Rx and were heated at 60°C for 1 hour, dipped in Xylene and mounted using EcoMount Mounting Medium (Biocare Medical, CA, USA. Cat No. EM897L).

| Probe Name | Catalogue Number |
| --- | --- |
| Hs HCAR2-1ZZ-st-C1 | 1137558-C1 |
| Hs HCAR3-1ZZ-st-C1 | 1155428-C1 |
| BA-Hs-PPIB-3zz | 701038 |
| BA-DapB-3zz | 701018 |

### Generation and characterisation of *Hcar2* p.R308C mice

#### Generation of the Hcar2 p.R308C mice

The R308C mutation was introduced into the mouse Hcar2 gene by the in-house transgenic facility utilizing CRISPR/Cas9 technology, as previously described^54^. Briefly, an sgRNA (5ʹ-AAGGCAGCGGTTGATACACG) that targets the R308C position in the genome was complexed with Cas9 protein and electroporated into C57BL/6NJ 1-cell embryos together with an oligo donor containing the missense mutation (CGC > TGT, underlined), 5ʹ-TTCTTCTCCACGTGTATCAACTGTTGCCTTCGAAAGAAAACATTGGGTGAACCCGATAAT. Embryos were then transferred to pseudo-pregnant females to generate pups. Potential founders were genotyped by genomic PCR (primers: 5ʹ-GCTCAGGCAGGATCATCTGG and 5ʹ-AAACTGCTCACTGCACCACG) followed by Sanger sequencing. Selected founders carrying the R308C mutation were bred with wildtype C57BL/6NJ ((JAX#005304) mice for germline transmission. Mice were bred heterozygote to heterozygote, and littermates were used for all experiments. All procedures performed on animals were in accordance with regulations and established guidelines and were reviewed and approved by an Institutional Animal Care and Use Committee or through an ethical review process.

#### Body weight and composition

Male mice homozygous for HCAR2 WT and HCAR2 R308C were group housed at room temperature and fed a standard chow diet (Inotiv Teklad 2916). Body weights were measured weekly from 2 months to 5 months of age, and every 2-4 weeks thereafter until 40-weeks of age. Body fat mass and lean mass were evaluated by EchoMRI at 5 months and 9 months of age. MicroCT was performed at 6 months of age using a Bruker Skyscan 1276 CT; briefly, a full-body CT-scan was performed, and an auto-segmentation tool was used to quantify subcutaneous and visceral adipose tissue volume based on CT image grayscale values. Tissue weights were collected at necropsy following a 2-hour fast at 12-months of age.

#### Insulin and oral glucose tolerance tests (ITT and OGTT)

For the ITT, 9-month-old mice were fasted for 4 hours and administered an intraperitoneal dose of insulin (Humulin) in saline at 0.75U/kg body weight. Blood glucose was collected via tail snip using an AlphaTrak2 glucometer (Zoetis) at baseline, 15 min, 30 min, 45 min, 60 min, 90 min, and 120 min after dosing. For the OGTT, mice were fasted for 6 hours prior to dosing with a glucose solution. Mice received 2g glucose/kg body weight via oral gavage. Blood glucose was collected via tail snip using an AlphaTrak2 glucometer (Zoetis) at baseline, 15 min, 30 min, 60 min, 90 min, and 120 min after dosing. Data for ITT is presented as a percentage of baseline glucose. Data for OGTT is presented as raw glucose values. AUC was calculated using the trapezoidal rule.

#### Glucose stimulated insulin secretion

Mice were fasted for 6 hours prior to dosing with a glucose solution. Mice were dosed with 2g glucose/kg body weight via oral gavage. Blood was collected via tail snip at baseline, 15 min, 30 min, and 120 min after dosing. Blood was centrifuged to collect plasma. Plasma insulin was then measured using the Crystal Chem Ultra-Sensitive Mouse Insulin ELISA Kit, 2nd Gen (Crystal Chem Cat# 62100) according to manufacturer’s instructions.

#### Fasting re-feed study

Mice were fasted overnight. In the morning, blood was collected via tail snip. Following fasting blood collection, chow diet was returned to the mice for the re-feed period. Two hours after returning the food, another blood sample was collected via tail snip. Blood was then centrifuged to collect plasma. Plasma NEFA levels were measured using the Fujifilm NEFA-HR (2) kit (Fisher Scientific NC9517308, NC9517309, NC9517310, and NC9517311).

#### Mouse plasma analysis

Liver enzymes were assayed using Siemens ADVIA Chemistry kits for ALT (SMN 10309500) and AST (SMN 10309496) on a Siemens Advia Chemistry XPT Clinical Analyzer. Cholesterol and TAG were measured by HPLC.

#### Hepatic TAG

Whole frozen livers were pulverized on dry ice and approximately 50mg was weighed and transferred into an Eppendorf tube with 500µL of lysis buffer (140mM NaCl, 50mM Tris pH 7.4, 0.2% Triton X-100). Samples were homogenized using a Tissuelyzer. Homogenate was diluted in PBS, transferred to a 96-well plate, and incubated with a total concentration of 0.5% deoxycholate for 5 minutes at 37°C. Infinity Triglyceride Reagent (Thermo Scientific TR22421) was added to each well and incubated for 20 minutes at 37°C. The plate was read at 500nm to determine TAG content, which was then normalized to tissue weight.

#### Ex vivo lipolysis

eWAT fad pads isolated from male mice expressing HCAR2-WT or HCAR2-p.R308C were rinsed in PBS and cut into 50mg pieces. Tissue pieces were suspended in lipolysis assay buffer (KRBH+0.2% fatty acid free-BSA (Roche 03117057001)) in a 96-well plate and equilibrated with shaking for 2 hours at 37°C. The tissue was then treated with 10mM BHB or control assay buffer for 20 minutes. The BHB solution was made fresh the day of the assay by suspending BHB (Sigma-Aldrich 166898) in lipolysis assay buffer and neutralizing to pH 7.4 with NaOH. Following stimulation, the tissue was then transferred into fresh assay buffer for 1 hour at 37°C with shaking. Supernatant NEFA levels were measured using the Fujifilm NEFA-HR (2) kit (Fisher Scientific NC9517308, NC9517309, NC9517310, and NC9517311) and normalized to tissue weight.

### In vitro characterisation of *HCAR2* p.R311C

#### BRET-based GPCR profiling

BRET-based biosensor studies were conducted using the bioSens-All® platform as previously described^55^. Briefly: HEK293 cells maintained in Dulbecco’s Modified Eagle Medium (Wisent #319-015-CL) supplemented with 1% penicillin-streptomycin (Wisent #450-201-EL) and 5% fetal bovine serum (Wisent #090150) were co-transfected with either *HCAR2* WT or *HCAR2* p.R311C and one of the following bioSensAll® assays: Gαi1, β-arrestin2+GRK2, or Rec-EndoE (early endosome). Following transfection using PEI reagent (polyethylenimine 25 kDa linear, PolyScience #23966), cells were plated at a density of 35K per well in opaque 96-well plates (Greiner #655083). At 48 hours post-transfection, culture medium was aspirated and replaced with 100 μl of Hank’s Balanced Salt Solution buffer (HBSS) (Wisent #319-067CL) per well. 10 μl of 20 μM e-Coelenterazine Prolume Purple (Methoxy e-CTZ) (Nanolight #369) was added to each well for a final concentration of 2 μM. One of six compounds was added to each well using the HP D300 digital dispenser (Tecan): Nicotinic acid, β-hydroxybutyrate, GSK-256073, MK-1903, MK-0354, or monomethyl fumarate). All compounds were assayed at 12 concentrations per biosensor. Cells were incubated with the compounds at room temperature for 10 minutes (for G-protein activation and β-arrestin recruitment) and at 37°C for 30 minutes for receptor internalization and trafficking. BRET readings were collected with a 0.02sec integration time on a Spark plate reader (Tecan Group Ltd.; filters 440 nm/360 nm, 575 nm/505 nm). BRET signals were determined by calculating the ratio of light emitted by GFP-acceptor (515nm) over light emitted by luciferase-donor (400 nm). Resulting dose-response curves were fitted using the four-parameter logistic non-linear regression model. All data is presented as percent nicotinic acid maximum.

#### Generation and culture of CHO cells

Human HCAR2 WT and HCAR2 p.R11C cDNA were synthesized and subcloned into pcDNA5/FRT/TO (Thermo Fisher, V652020). The pcDNA5/FRT/TO/ human HCAR2 plasmids were then transfected into the Flp-In™-CHO Cell Line (Thermo Fisher, R75807) and doxycycline-inducible clonal cell lines were selected (CHO TREx HCAR2 WT, or CHO TREx HCAR2 R311C). Sequences encoding HiBit-tagged versions of the wild-type human HCAR2 WT and R311C were synthesized and cloned into pcDNA3.1. The HiBit (VSGWRLFKKIS) tag was sequentially included after the N-terminal initiator methionine. The nucleotide sequences of all receptor constructs were confirmed by automated DNA sequencing. Unless otherwise specified, all studies were performed without the addition of doxycycline (“low expression” conditions). Cells were maintained in complete media containing DMEM-F12 (Gibco #10565), 10% FBS (Gibco A5670801), 1% Glutamax (Gibco 35050061), 15ug/mL Blasticidin (Gibco R21001), 700 µg/mL Hygromycin (Gibco 10687010), and 100 µg/mL Normocin (Invivogen #ant-nr-2).

#### HiBiT assa

CHO TrEX Hibit-HCAR2 WT or R311C were plated for 48 hours in complete media (described above). The day of the assay, cells were dissociated, counted, and resuspended in assay buffer (phenol-free DMEM (Gibco 31053036) with 2% FBS). Cells were then dispensed in a non-binding 384-well plate at a density of 7.5K cells/well. HiBiT quantification was performed using the Nano-Glo® HiBiT Extracellular Detection System (Promega N2421) for surface expression and the Nano-Glo® HiBiT Lytic Detection System (Promega N3040) for total expression. Luminescence was measured with an EnVision plate reader to determine surface expression. The raw data was normalized to percent WT maximum for both surface and total expression.

#### RTqPCR

RNA was isolated from CHO TrEX Hibit-HCAR2 WT or R311C cell lines using an RNEasy kit (QIAGEN #74104) with on-column DNA digestion (QIAGEN #79254). Reverse transcription was performed using the Applied Biosciences High-Capacity cDNA Reverse Transcription Kit (ThermoFisher #4368814). RTqPCR was performed by Applied Biosystems TaqMan Real-Time PCR assays using probes for *HCAR2* (Hs02341584_s1) and *HPRT* (Hs01003267_m1). Relative gene expression was calculated using ΔΔCT and normalized to *HPRT*.

#### Western blot

CHO TrEX Hibit-HCAR2 WT or R311C were plated overnight in complete media with control or 1 µg/uL doxycycline to induce high expression of HCAR2. Cells were lysed in RIPA buffer (ThermoScientific 89900) + Pierce protease/phosphatase inhibitors (ThermoFisher A32961) and protein concentration was quantified using a Pierce BCA kit (ThermoFisher 23225). Cell lysates were diluted with 4X NuPAGE LDS Sample Buffer (Invitrogen NP0007) and 10X NuPAGE Sample Reducing Agent (Invitrogen NP0009). Western blot was run by standard-SDS PAGE using a BioRad Criterion gel system. Membranes were incubated in primary antibodies overnight (Anti-β-actin (Cell Signalling #4967), anti-Hibit antibody (Promega N7200), or anti-HCAR2/GPR109a antibody (Invitrogen/Thermo PA5-90579)). Membranes were washed and incubated in secondary antibodies (Anti-rabbit IgG, HRP-linked Antibody (Cell Signaling #7074) or Anti-mouse IgG, HRP-linked Antibody (Cell Signaling #7076)) before imaging on the BioRad Chemi Doc system.

#### 3H niacin adipose membrane binding

iWAT fat pads were harvested from female HCAR2 WT and p.R308C mice, minced, and suspended in homogenization buffer (20mM Tris-HCl, pH 7.5, 1mM EDTA, 1 mM EGTA, 250 mM sucrose). Minced tissue was homogenized using a Polytron PT-MR 3100 homogenizer and centrifuged at 2000 x *g* for 10 minutes to remove the fat layer. Homogenate was then ultracentrifuged at 45,000 x *g* for 45 minutes. The pelleted membranes were resuspended in homogenization buffer + protease inhibitors (ThermoFisher A32961) and dispersed using a dounce homogenizer. Membrane homogenate was quantified using a Pierce BCA kit (ThermoFisher 23225). A similar method was used to isolate membranes from CHO TrEX Hibit-HCAR2 WT cells to serve as a control. Membrane was suspended in assay buffer (HBSS with 10 mM HEPES and 1% BSA) and plated at a concentration of 60 µg/well (mouse adipose) or 3.3 ug/well (CHO control cells). 60nM [3H]-Niacin (American Radiolabeled Chemicals, #ART 0689) was added to evaluate specific binding and 10uM cold niacin was added to evaluate nonspecific binding (Sigma PHR1276). After a 2-hour incubation at room temperature, membrane was harvested and transferred to a glass fiber filter plate (Revvity #6055690) using a TomTec harvester. After drying the filter plate overnight at room temperature, Ultima Gold scintillation fluid (Revvity #6013681) was added and CPM were quantified using a Microbetta Scintillation Counter. Nonspecific binding was subtracted from total binding to calculate specific binding. All data was normalized to percent binding of [3H]-Niacin to the CHO TrEX Hibit-HCAR2 WT control.

## Data availa1bility

Data from Fenland, EPIC-Norfolk and ELY studies can be requested by bona fide researchers for specified scientific purposes via the following study websites:

Fenland: https://www.mrc-epid.cam.ac.uk/research/studies/fenland

EPIC-Norfolk: https://www.mrc-epid.cam.ac.uk/research/studies/epic-norfolk

ELY: https://www.mrc-epid.cam.ac.uk/research/studies/ely

Data will either be shared through an institutional data sharing agreement or arrangements will be made for analyses to be conducted remotely without the necessity for data transfer.

Oxford Biobank is accessible via the following website upon application: https://www.oxfordbiobank.org.uk/for-researchers-2/

All data used in the genetic association analyses are available from the UKBB upon application (https://www.ukbiobank.ac.uk).

## Acknowledgements

We thank the participants and investigators in the UK Biobank (project numbers 32974 and 26041), Fenland, Epic-Norfolk, Ely, Oxford Biobank and the FPLD1 studies who made this work possible. S.O., B.Y.H.L. and K.R. are supported by the MRC Metabolic Diseases Unit (MC_UU_00039/1). S.L. is supported by a Wellcome Trust Clinical PhD Fellowship (225479/Z/22). HCAR2 and HCAR3 genotyping was performed at the Institute of Metabolic Science Genomics and Bioinformatics Core supported by the MRC (MC_UU_00039), the Wellcome Discovery Research Platform (226800/Z/22/Z). F.R.D., A.W., R.J., V.K., M.A., K.K.O., J.R.B.P. and N.J.W are supported by the MRC (MC_UU_00006/1, MC_UU_00006/2). D.A-V is funded by the Instituto de Salud Carlos III PI22/00514 and and co-funded by the European Union and Xunta de Galicia ED431C 2024/11. D.B.S. is supported by the Wellcome Trust (WT 219417), the MRC (MR/X00970X/1), and The National Institute for Health Research (NIHR) Cambridge Biomedical Research Centre and NIHR Rare Disease Translational Research Collaboration. D.J.F. is supported by a Medical Research Council career development award (MR/S007091/1). JAT is supported by BBSRC Project Grant (BB/S017593/1). We’d also like to extend our appreciation to Pfizer colleagues who provided *in vivo* technical support for the development and characterization of the HCAR2 p.R308C mouse model, including Youngwook Ahn, Danielle Archambault, Mackenzie Marshall, Jeffrey Morin, Rachel Poskanzer, and Alexis White.

## References

1. Reaven, G.M. (1993). Role of insulin resistance in human disease (syndrome X): an expanded definition. Annu Rev Med 44, 121–131. 10.1146/annurev.me.44.020193.001005.

2. Graham, S.E., Clarke, S.L., Wu, K.H., Kanoni, S., Zajac, G.J.M., Ramdas, S., Surakka, I., Ntalla, I., Vedantam, S., Winkler, T.W., et al. (2021). The power of genetic diversity in genome-wide association studies of lipids. Nature 600, 675–679. 10.1038/s41586-021-04064-3.

3. Koprulu, M., Zhao, Y., Wheeler, E., Dong, L., Rocha, N., Li, C., Griffin, J.D., Patel, S., Van de Streek, M., Glastonbury, C.A., et al. (2022). Identification of Rare Loss-of-Function Genetic Variation Regulating Body Fat Distribution. J Clin Endocrinol Metab 107, 1065–1077. 10.1210/clinem/dgab877.

4. Lotta, L.A., Gulati, P., Day, F.R., Payne, F., Ongen, H., van de Bunt, M., Gaulton, K.J., Eicher, J.D., Sharp, S.J., Luan, J., et al. (2017). Integrative genomic analysis implicates limited peripheral adipose storage capacity in the pathogenesis of human insulin resistance. Nat Genet 49, 17–26. 10.1038/ng.3714.

5. Pulit, S.L., Stoneman, C., Morris, A.P., Wood, A.R., Glastonbury, C.A., Tyrrell, J., Yengo, L., Ferreira, T., Marouli, E., Ji, Y., et al. (2019). Meta-analysis of genome-wide association studies for body fat distribution in 694 649 individuals of European ancestry. Hum Mol Genet 28, 166–174. 10.1093/hmg/ddy327.

6. Williamson, A., Norris, D.M., Yin, X., Broadaway, K.A., Moxley, A.H., Vadlamudi, S., Wilson, E.P., Jackson, A.U., Ahuja, V., Andersen, M.K., et al. (2023). Genome-wide association study and functional characterization identifies candidate genes for insulin-stimulated glucose uptake. Nat Genet 55, 973–983. 10.1038/s41588-023-01408-9.

7. Sakers, A., De Siqueira, M.K., Seale, P., and Villanueva, C.J. (2022). Adipose-tissue plasticity in health and disease. Cell 185, 419–446. 10.1016/j.cell.2021.12.016.

8. Gandotra, S., Le Dour, C., Bottomley, W., Cervera, P., Giral, P., Reznik, Y., Charpentier, G., Auclair, M., Delepine, M., Barroso, I., et al. (2011). Perilipin deficiency and autosomal dominant partial lipodystrophy. N Engl J Med 364, 740–748. 10.1056/NEJMoa1007487.

9. Rubio-Cabezas, O., Puri, V., Murano, I., Saudek, V., Semple, R.K., Dash, S., Hyden, C.S., Bottomley, W., Vigouroux, C., Magre, J., et al. (2009). Partial lipodystrophy and insulin resistant diabetes in a patient with a homozygous nonsense mutation in CIDEC. EMBO Mol Med 1, 280–287. 10.1002/emmm.200900037.

10. Deaton, A.M., Dubey, A., Ward, L.D., Dornbos, P., Flannick, J., Consortium, A.-T.D.G., Yee, E., Ticau, S., Noetzli, L., Parker, M.M., et al. (2022). Rare loss of function variants in the hepatokine gene INHBE protect from abdominal obesity. Nat Commun 13, 4319. 10.1038/s41467-022-31757-8.

11. Cai, T.Q., Ren, N., Jin, L., Cheng, K., Kash, S., Chen, R., Wright, S.D., Taggart, A.K., and Waters, M.G. (2008). Role of GPR81 in lactate-mediated reduction of adipose lipolysis. Biochem Biophys Res Commun 377, 987–991. 10.1016/j.bbrc.2008.10.088.

12. Liu, C., Wu, J., Zhu, J., Kuei, C., Yu, J., Shelton, J., Sutton, S.W., Li, X., Yun, S.J., Mirzadegan, T., et al. (2009). Lactate inhibits lipolysis in fat cells through activation of an orphan G-protein-coupled receptor, GPR81. J Biol Chem 284, 2811–2822. 10.1074/jbc.M806409200.

13. Taggart, A.K., Kero, J., Gan, X., Cai, T.Q., Cheng, K., Ippolito, M., Ren, N., Kaplan, R., Wu, K., Wu, T.J., et al. (2005). (D)-beta-Hydroxybutyrate inhibits adipocyte lipolysis via the nicotinic acid receptor PUMA-G. J Biol Chem 280, 26649–26652. 10.1074/jbc.C500213200.

14. Ahmed, K., Tunaru, S., Langhans, C.D., Hanson, J., Michalski, C.W., Kolker, S., Jones, P.M., Okun, J.G., and Offermanns, S. (2009). Deorphanization of GPR109B as a receptor for the beta-oxidation intermediate 3-OH-octanoic acid and its role in the regulation of lipolysis. J Biol Chem 284, 21928–21933. 10.1074/jbc.M109.019455.

15. Ahmed, K., Tunaru, S., and Offermanns, S. (2009). GPR109A, GPR109B and GPR81, a family of hydroxy-carboxylic acid receptors. Trends Pharmacol Sci 30, 557–562. 10.1016/j.tips.2009.09.001.

16. Justice, A.E., Karaderi, T., Highland, H.M., Young, K.L., Graff, M., Lu, Y., Turcot, V., Auer, P.L., Fine, R.S., Guo, X., et al. (2019). Protein-coding variants implicate novel genes related to lipid homeostasis contributing to body-fat distribution. Nat Genet 51, 452–469. 10.1038/s41588-018-0334-2.

17. Liu, D.J., Peloso, G.M., Yu, H., Butterworth, A.S., Wang, X., Mahajan, A., Saleheen, D., Emdin, C., Alam, D., Alves, A.C., et al. (2017). Exome-wide association study of plasma lipids in >300,000 individuals. Nat Genet 49, 1758–1766. 10.1038/ng.3977.

18. Zellner, C., Pullinger, C.R., Aouizerat, B.E., Frost, P.H., Kwok, P.Y., Malloy, M.J., and Kane, J.P. (2005). Variations in human HM74 (GPR109B) and HM74A (GPR109A) niacin receptors. Hum Mutat 25, 18–21. 10.1002/humu.20121.

19. Tuteja, S., Wang, L., Dunbar, R.L., Chen, J., DerOhannessian, S., Marcovina, S.M., Elam, M., Lader, E., and Rader, D.J. (2017). Genetic coding variants in the niacin receptor, hydroxyl-carboxylic acid receptor 2, and response to niacin therapy. Pharmacogenet Genomics 27, 285–293. 10.1097/FPC.0000000000000289.

20. Lotta, L.A., Scott, R.A., Sharp, S.J., Burgess, S., Luan, J., Tillin, T., Schmidt, A.F., Imamura, F., Stewart, I.D., Perry, J.R., et al. (2016). Genetic Predisposition to an Impaired Metabolism of the Branched-Chain Amino Acids and Risk of Type 2 Diabetes: A Mendelian Randomisation Analysis. PLoS Med 13, e1002179. 10.1371/journal.pmed.1002179.

21. Forouhi, N.G., Luan, J., Hennings, S., and Wareham, N.J. (2007). Incidence of Type 2 diabetes in England and its association with baseline impaired fasting glucose: the Ely study 1990-2000. Diabet Med 24, 200–207. 10.1111/j.1464-5491.2007.02068.x.

22. Day, N., Oakes, S., Luben, R., Khaw, K.T., Bingham, S., Welch, A., and Wareham, N. (1999). EPIC-Norfolk: study design and characteristics of the cohort. European Prospective Investigation of Cancer. Br J Cancer 80 Suppl 1, 95–103.

23. Karpe, F., Vasan, S.K., Humphreys, S.M., Miller, J., Cheeseman, J., Dennis, A.L., and Neville, M.J. (2018). Cohort Profile: The Oxford Biobank. Int J Epidemiol 47, 21–21g. 10.1093/ije/dyx132.

24. Hung, J., McQuillan, B.M., Thompson, P.L., and Beilby, J.P. (2008). Circulating adiponectin levels associate with inflammatory markers, insulin resistance and metabolic syndrome independent of obesity. Int J Obes (Lond) 32, 772–779. 10.1038/sj.ijo.0803793.

25. Segerstolpe, A., Palasantza, A., Eliasson, P., Andersson, E.M., Andreasson, A.C., Sun, X., Picelli, S., Sabirsh, A., Clausen, M., Bjursell, M.K., et al. (2016). Single-Cell Transcriptome Profiling of Human Pancreatic Islets in Health and Type 2 Diabetes. Cell Metab 24, 593–607. 10.1016/j.cmet.2016.08.020.

26. Guillin-Amarelle, C., Sanchez-Iglesias, S., Castro-Pais, A., Rodriguez-Canete, L., Ordonez-Mayan, L., Pazos, M., Gonzalez-Mendez, B., Rodriguez-Garcia, S., Casanueva, F.F., Fernandez-Marmiesse, A., and Araujo-Vilar, D. (2016). Type 1 familial partial lipodystrophy: understanding the Kobberling syndrome. Endocrine 54, 411–421. 10.1007/s12020-016-1002-x.

27. Jadeja, R.N., Jones, M.A., Fromal, O., Powell, F.L., Khurana, S., Singh, N., and Martin, P.M. (2019). Loss of GPR109A/HCAR2 induces aging-associated hepatic steatosis. Aging (Albany NY) 11, 386–400. 10.18632/aging.101743.

28. Ye, L., Cao, Z., Lai, X., Wang, W., Guo, Z., Yan, L., Wang, Y., Shi, Y., and Zhou, N. (2019). Niacin fine-tunes energy homeostasis through canonical GPR109A signaling. FASEB J 33, 4765–4779. 10.1096/fj.201801951R.

29. Arner, P., Andersson, D.P., Backdahl, J., Dahlman, I., and Ryden, M. (2018). Weight Gain and Impaired Glucose Metabolism in Women Are Predicted by Inefficient Subcutaneous Fat Cell Lipolysis. Cell Metab 28, 45–54 e43. 10.1016/j.cmet.2018.05.004.

30. Puchalska, P., and Crawford, P.A. (2021). Metabolic and Signaling Roles of Ketone Bodies in Health and Disease. Annu Rev Nutr 41, 49–77. 10.1146/annurev-nutr-111120-111518.

31. Pietrzak, D., Kasperek, K., Rekawek, P., and Piatkowska-Chmiel, I. (2022). The Therapeutic Role of Ketogenic Diet in Neurological Disorders. Nutrients 14. 10.3390/nu14091952.

32. Zhu, H., Bi, D., Zhang, Y., Kong, C., Du, J., Wu, X., Wei, Q., and Qin, H. (2022). Ketogenic diet for human diseases: the underlying mechanisms and potential for clinical implementations. Signal Transduct Target Ther 7, 11. 10.1038/s41392-021-00831-w.

33. Myette-Cote, E., Caldwell, H.G., Ainslie, P.N., Clarke, K., and Little, J.P. (2019). A ketone monoester drink reduces the glycemic response to an oral glucose challenge in individuals with obesity: a randomized trial. Am J Clin Nutr 110, 1491–1501. 10.1093/ajcn/nqz232.

34. Owen, O.E., Felig, P., Morgan, A.P., Wahren, J., and Cahill, G.F., Jr. (1969). Liver and kidney metabolism during prolonged starvation. J Clin Invest 48, 574–583. 10.1172/JCI106016.

35. Grunenwald, A., Peliconi, J., Zarantonello, A., Dimitrov, J.D., Roumenina, L.T., and Merle, N.S. (2025). HCAR2 is a novel receptor for heme. Blood Adv. 10.1182/bloodadvances.2025016197.

36. Brownsey, R.W., Boone, A.N., Elliott, J.E., Kulpa, J.E., and Lee, W.M. (2006). Regulation of acetyl-CoA carboxylase. Biochem Soc Trans 34, 223–227. 10.1042/BST20060223.

37. Vice, E., Privette, J.D., Hickner, R.C., and Barakat, H.A. (2005). Ketone body metabolism in lean and obese women. Metabolism 54, 1542–1545. 10.1016/j.metabol.2005.05.023.

38. Rochford, J.J. (2022). When Adipose Tissue Lets You Down: Understanding the Functions of Genes Disrupted in Lipodystrophy. Diabetes 71, 589–598. 10.2337/dbi21-0006.

39. Gabe, M.B.N., van der Velden, W.J.C., Gadgaard, S., Smit, F.X., Hartmann, B., Brauner-Osborne, H., and Rosenkilde, M.M. (2020). Enhanced agonist residence time, internalization rate and signalling of the GIP receptor variant [E354Q] facilitate receptor desensitization and long-term impairment of the GIP system. Basic Clin Pharmacol Toxicol 126 *Suppl 6*, 122–132. 10.1111/bcpt.13289.

40. Mohammad, S., Patel, R.T., Bruno, J., Panhwar, M.S., Wen, J., and McGraw, T.E. (2014). A naturally occurring GIP receptor variant undergoes enhanced agonist-induced desensitization, which impairs GIP control of adipose insulin sensitivity. Mol Cell Biol 34, 3618–3629. 10.1128/MCB.00256-14.

41. Li, G., Zhou, Q., Yu, Y., Chen, L., Shi, Y., Luo, J., Benovic, J., Lu, J., and Zhou, N. (2012). Identification and characterization of distinct C-terminal domains of the human hydroxycarboxylic acid receptor-2 that are essential for receptor export, constitutive activity, desensitization, and internalization. Mol Pharmacol 82, 1150–1161. 10.1124/mol.112.081307.

42. Anwar, M.U., and van der Goot, F.G. (2023). Refining S-acylation: Structure, regulation, dynamics, and therapeutic implications. J Cell Biol 222. 10.1083/jcb.202307103.

43. Jansen, M., and Beaumelle, B. (2022). How palmitoylation affects trafficking and signaling of membrane receptors. Biol Cell 114, 61–72. 10.1111/boc.202100052.

44. (!!! INVALID CITATION !!!, which could contribute to the reduction in HCAR2 p.R311C a).

45. Soga, T., Kamohara, M., Takasaki, J., Matsumoto, S., Saito, T., Ohishi, T., Hiyama, H., Matsuo, A., Matsushime, H., and Furuichi, K. (2003). Molecular identification of nicotinic acid receptor. Biochem Biophys Res Commun 303, 364–369. 10.1016/s0006-291x(03)00342-5.

46. Tunaru, S., Kero, J., Schaub, A., Wufka, C., Blaukat, A., Pfeffer, K., and Offermanns, S. (2003). PUMA-G and HM74 are receptors for nicotinic acid and mediate its anti-lipolytic effect. Nat Med 9, 352–355. 10.1038/nm824.

47. Wise, A., Foord, S.M., Fraser, N.J., Barnes, A.A., Elshourbagy, N., Eilert, M., Ignar, D.M., Murdock, P.R., Steplewski, K., Green, A., et al. (2003). Molecular identification of high and low affinity receptors for nicotinic acid. J Biol Chem 278, 9869–9874. 10.1074/jbc.M210695200.

48. Dobbins, R., Byerly, R., Gaddy, R., Gao, F., Mahar, K., Napolitano, A., Ambery, P., and Le Monnier de Gouville, A.C. (2015). GSK256073 acutely regulates NEFA levels via HCA2 agonism but does not achieve durable glycaemic control in type 2 diabetes. A randomised trial. Eur J Pharmacol 755, 95–101. 10.1016/j.ejphar.2015.03.005.

49. Lai, E., Waters, M.G., Tata, J.R., Radziszewski, W., Perevozskaya, I., Zheng, W., Wenning, L., Connolly, D.T., Semple, G., Johnson-Levonas, A.O., et al. (2008). Effects of a niacin receptor partial agonist, MK-0354, on plasma free fatty acids, lipids, and cutaneous flushing in humans. J Clin Lipidol 2, 375–383. 10.1016/j.jacl.2008.08.445.

50. Hewitt, J., Walters, M., Padmanabhan, S., and Dawson, J. (2016). Cohort profile of the UK Biobank: diagnosis and characteristics of cerebrovascular disease. BMJ Open 6, e009161. 10.1136/bmjopen-2015-009161.

51. Gardner, E.J., Kentistou, K.A., Stankovic, S., Lockhart, S., Wheeler, E., Day, F.R., Kerrison, N.D., Wareham, N.J., Langenberg, C., O’Rahilly, S., et al. (2022). Damaging missense variants in IGF1R implicate a role for IGF-1 resistance in the etiology of type 2 diabetes. Cell Genom 2, None. 10.1016/j.xgen.2022.100208.

52. Powell, R.D.L.R., E.; Day, F.R.; Perry, J.R.B.; Griffin, S.J.; Forouhi, N.G.; Brage, S.; Wareham, N.J.; Langenberg, C.; Ong, K.K. (2020). Development and validation of total and regional body composition prediction equations from anthropometry and single frequency segmental bioelectrical impedance with DEXA. medRxiv. 10.1101/2020.12.16.20248330.

53. Cardona, A., Day, F.R., Perry, J.R.B., Loh, M., Chu, A.Y., Lehne, B., Paul, D.S., Lotta, L.A., Stewart, I.D., Kerrison, N.D., et al. (2019). Epigenome-Wide Association Study of Incident Type 2 Diabetes in a British Population: EPIC-Norfolk Study. Diabetes 68, 2315–2326. 10.2337/db18-0290.

54. Griffin, J.D., Buxton, J.M., Culver, J.A., Barnes, R., Jordan, E.A., White, A.R., Flaherty, S.E., Bernardo, B., Ross, T., Bence, K.K., and Birnbaum, M.J. (2023). Hepatic Activin E mediates liver-adipose inter-organ communication, suppressing adipose lipolysis in response to elevated serum fatty acids. Mol Metab 78, 101830. 10.1016/j.molmet.2023.101830.

55. Avet, C., Mancini, A., Breton, B., Le Gouill, C., Hauser, A.S., Normand, C., Kobayashi, H., Gross, F., Hogue, M., Lukasheva, V., et al. (2022). Effector membrane translocation biosensors reveal G protein and betaarrestin coupling profiles of 100 therapeutically relevant GPCRs. Elife 11. 10.7554/eLife.74101.

