## Supplementary Figures for "Genetic impairment of ketone body signalling is a prevalent contributor to human metabolic dysfunction"

Supplementary Figure 1

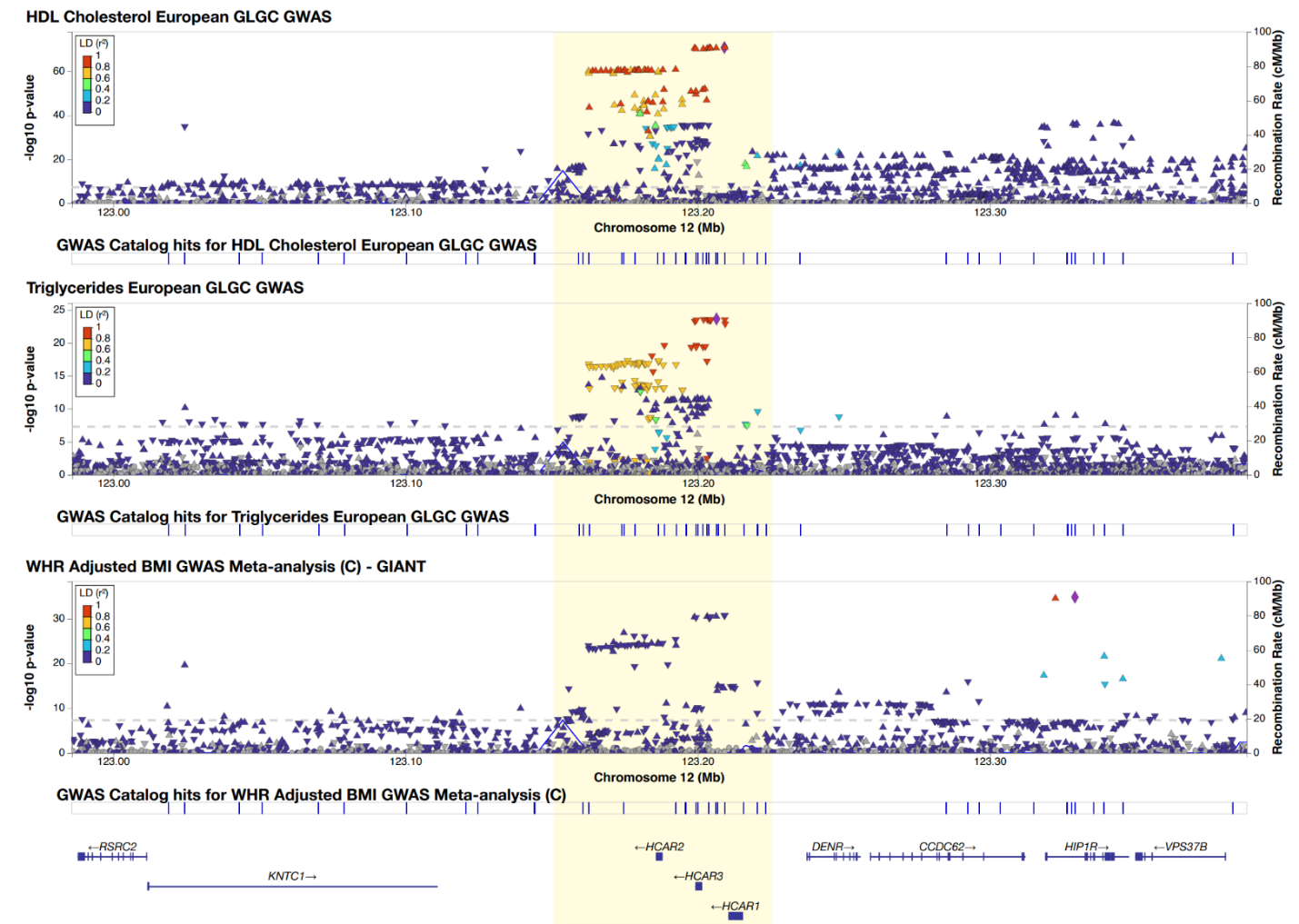

Locuszoom plots showing associations of the HCAR locus with HDL cholesterol and triglycerides (TAG) from the Global Lipids Genetics Corsortium (GLGC); and waist-hip ratio adjusted for BMI (WHRadjBMI) from the Genetic Investigation of ANthropomentric Traits (GIANT).

Supplementary Figure 2

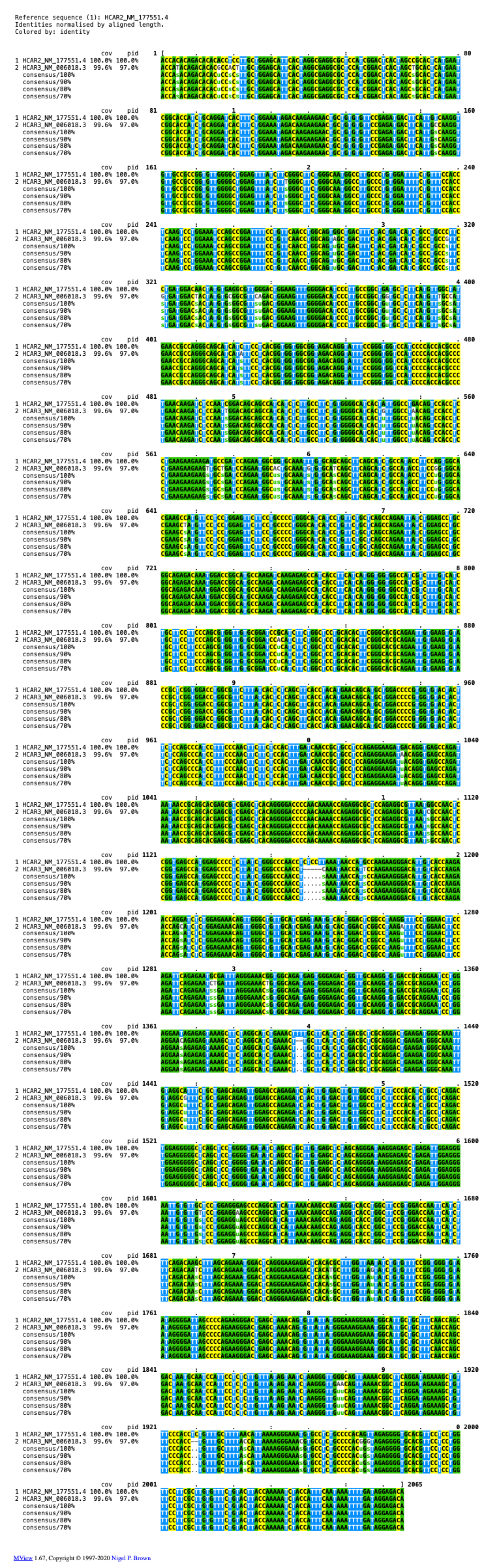

Pairwise clustal alignment of human *HCAR2* (NM_177551.4) and *HCAR3* (NM_006018.3) *c*DNA showing 97% sequence similarity between the two genes.

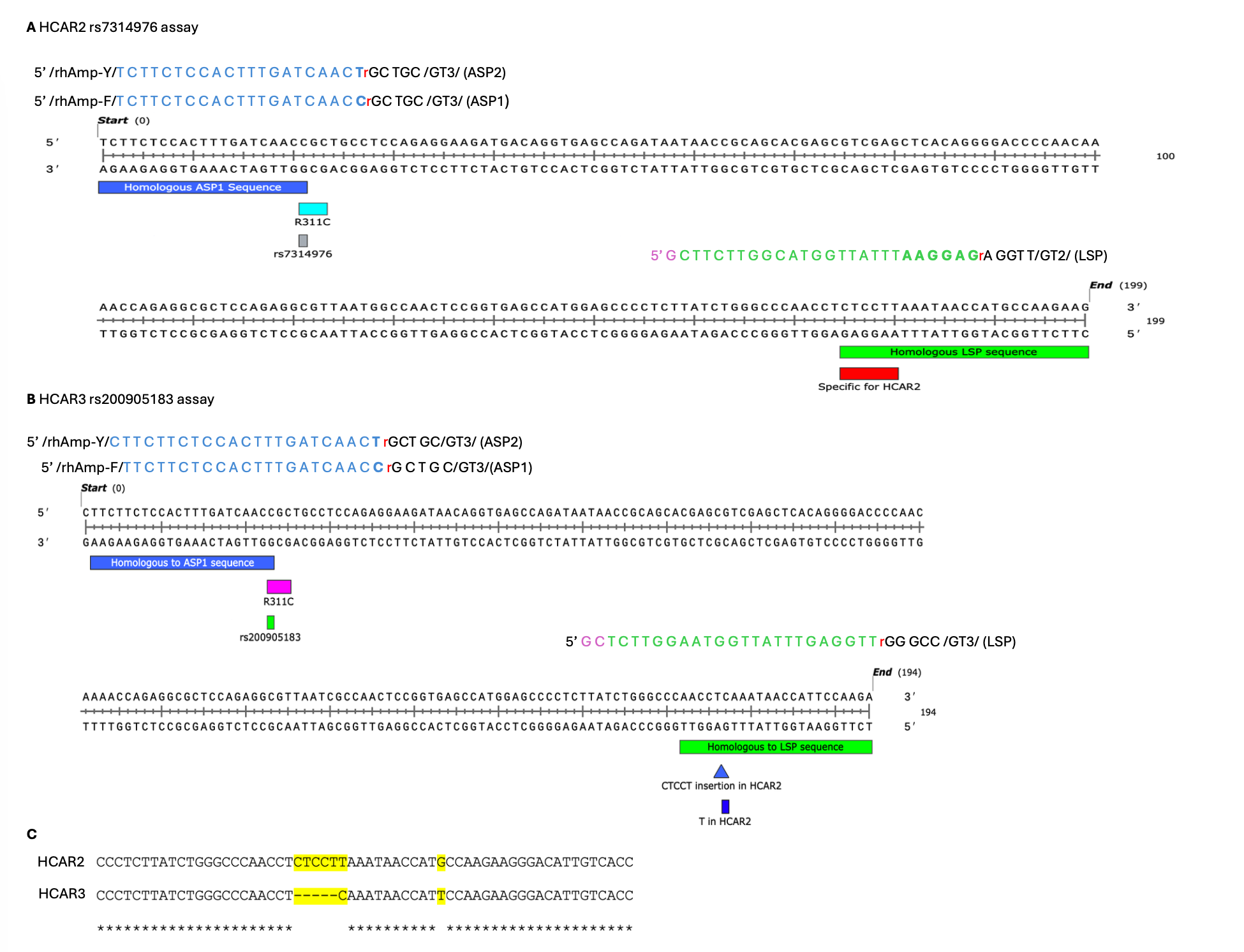
Supplementary Figure 3

Probe design for the quantitative PCR rhAMP genotyping assay. **(A) HCAR2 rs7314976 rhAmp™ SNP Genotyping Assay design.**

rhAmp™ SNP (IDT™ Integrated DNA Technologies) HCAR2 rs7314976 genotyping assay design number CD.GT.XFZV8105.1. rhAmp™ SNP genotyping assays contain three primers. Two allele specific primers (ASP1 wildtype allele and ASP2 for the alternative allele) and one locus specific primer (LSP). The nucleotide sequences in blue denotes the bases that are homologous to HCAR2 up to and includes the SNP allele in bold in the forward ASP1 and ASP2 primers. rhAmp-F and rhAmp-Y are proprietary designs (IDT) and are responsible for the allele detection via Fam for ASP1 and Yakima Yellow for ASP2. The nucleotide bases in green are homologous to HCAR2 and contain 6 bases “CTCCTT” in the forward strand, (green in bold in the reverse primer) that are highly HCAR2 specific and are not present in the homologus HCAR3 region, this has also been highlighted by the red box. All three primers contain an RNA base denoted by a “red r”. Bases 3’ of the RNA base are the blocking motif which prevents amplification continuing unless the primer hybridization is matched perfectly, then these bases are cleaved by the RNase H2 enzyme in the master mix and allowing the amplification to continue, producing a highly specific genotyping assay. The purple bases 5’ in the reverse LSP primer are there to assist the assay and this is again a proprietary design (IDT). (B) rhAmp™ SNP (IDT™ Integrated DNA Technologies) HCAR3 rs200905183 genotyping assay design number Hs.GT.rs200905183.A.1. Similarly this assay also consists of ASP1, ASP2 and LSP. The nucleotide sequences in blue denotes the bases that are homologous to HCAR3 up to and including the SNP allele in bold in the forward ASP1 Wildtype and ASP2 alternative forward primers. The nucleotide bases in green are highly homologous to HCAR3. The CTCCT insertion and the T bp in HCAR2 being absent here mean that this LSP sequence above specific for HCAR3 and not HCAR2. (C) Clustal Alignment of the HCAR2 versus HCAR3 gene sequence in the region where the Locus Specific Primers (LSP) for both the HCAR2 and HCAR3 assays have been designed.
Supplementary Figure 4

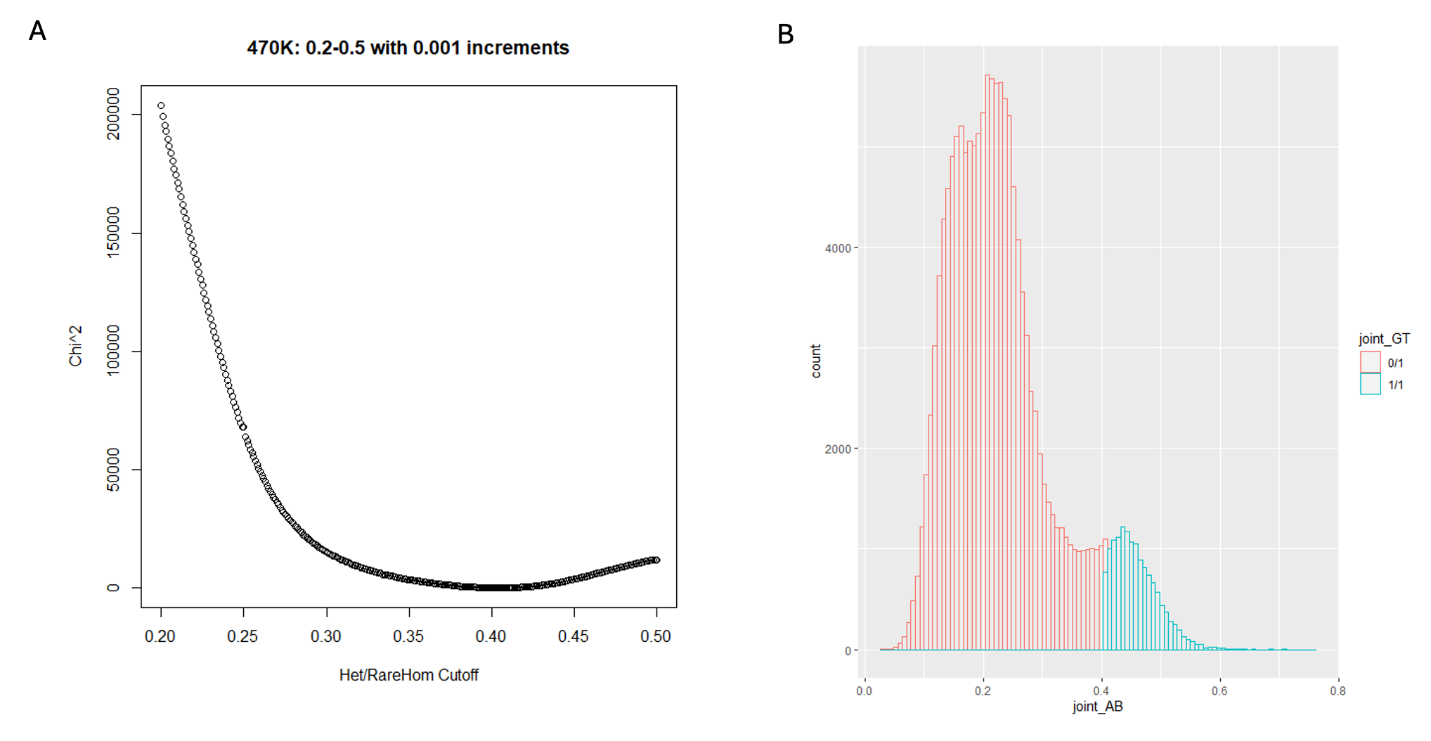

Genotyping call recalibration of *HCAR2 p.R311C (rs7314976 G>A)* in the UK Biobank. Both the reference and variant allele counts for both *HCAR2 (rs7314976)* and *HCAR3 (rs200905183)* sites were aggregated to form the new combined HCAR2/3 p.R311C. Next, a new variant allele balance (AB) cutoff for calling between hetero- or homozygous was determined by fitting genotyping counts to the Hardy Weinburg Equilibrium (HWE). (**A**) Scatterplot showing the HWE chi-square values with increasing AB cutoff from 0.2 to 0.5 with increments of 0.001. The AB resulting with the lowest chi-square was chosen as the homozygosity cutoff (0.41). (**B**) Histogram showing the new heterozygous (0/1, red) and alternate homozygous (1/1, turquoise) carriages of *HCAR2 p.R311C (rs7314976 G>A)* after recalibration.

Supplementary Figure 5

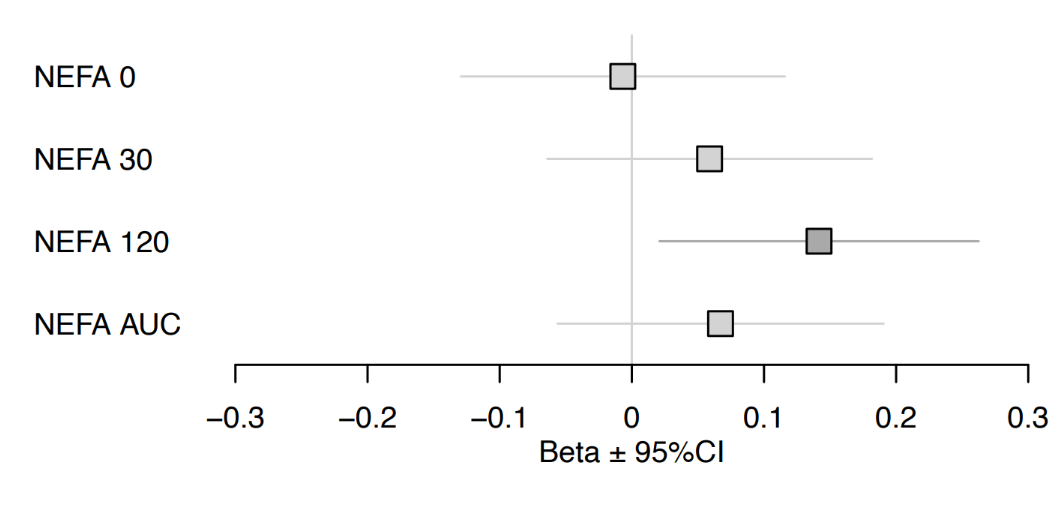

Association of HCAR2 p.R311C with plasma non-esterified fatty acids levels at 0, 30 and 120 minutes and area under the curve (AUC) after oral glucose tolerance test in the Ely Study (N=834, 823, 827 and 811 respectively)

Supplementary Figure 6

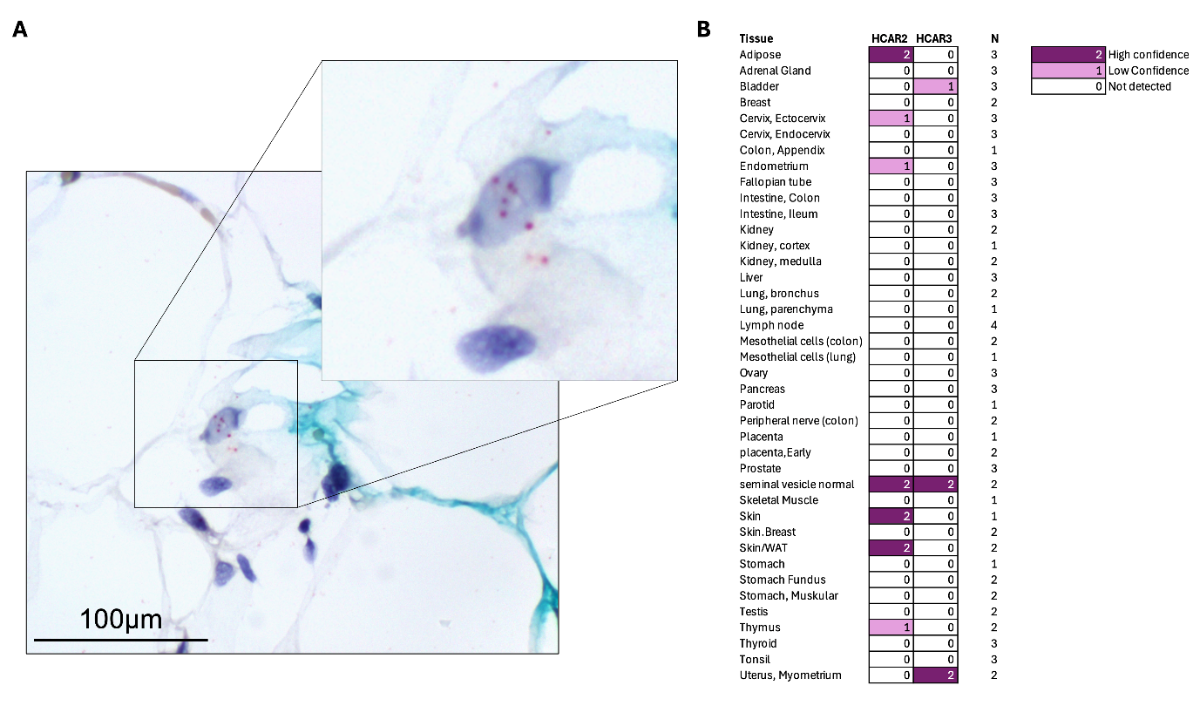

Tissue expression of *HCAR2* and *HCAR3* mRNA in human tissue by single-molecule in-situ hybridisation. **(A)** Brightfield microscopy image showing the expression of *HCAR2* mRNA punta (red) in human subcutaneous adipose tissue. **(B)** Expression summary of *HCAR2* and *HCAR3* in adipose and other tissues: 2=high-confidence expression; 1=low-confidence and 0=not detected.

Supplementary Figure 7

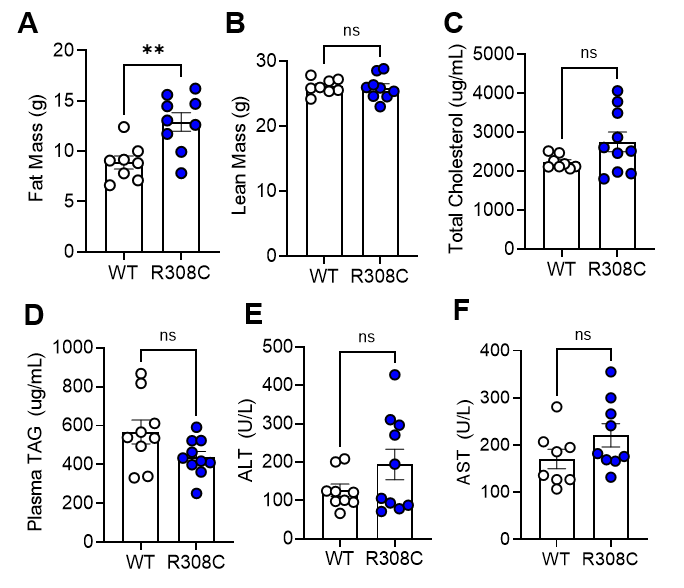

(A) Fat mass and (B) lean mass of WT and p.R308C mice at 9 months of age measured by EchoMRI; unpaired t-test. (C) Total cholesterol, (D) plasma TAG, (E) plasma ALT, and (F) plasma AST of WT and p.R308C mice at 12 months of age. For A-F, N=9 WT and 10 p.R308C males. Body weight and EchoMRI findings have been in repeated in an additional cohort. All values presented as mean ± SEM. *P < 0.05 and **P < 0.01.

Supplementary Figure 8

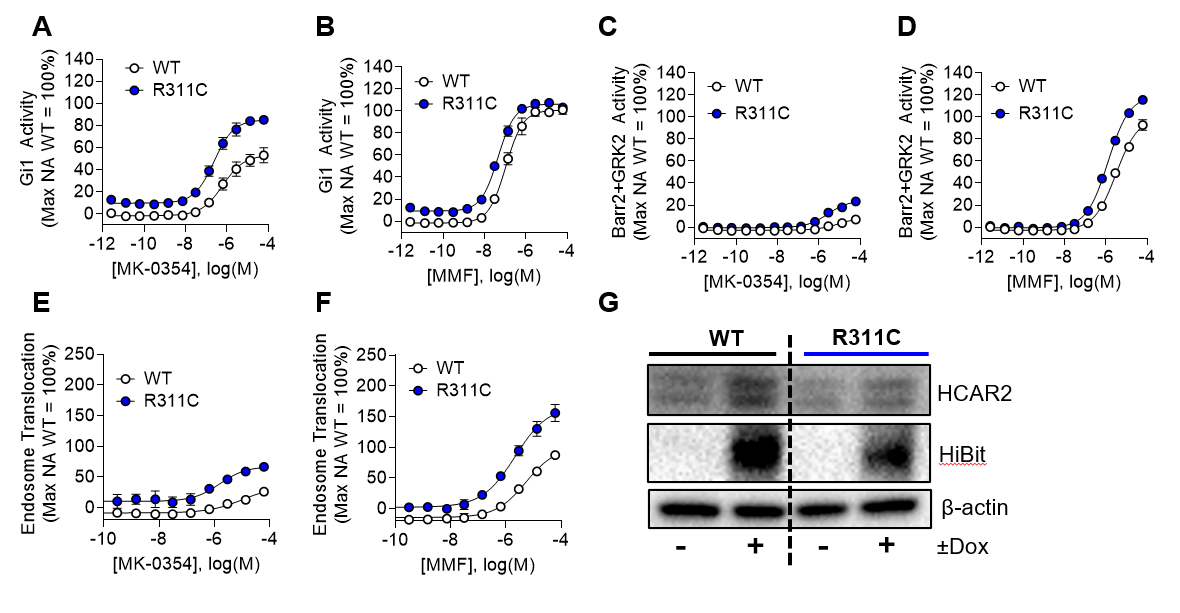

(A-B) Gi1 signalling activity of HEK 293 cells transiently transfected with *HCAR2-*WT or *HCAR2-*p.R311C. Cells were stimulated with (A) MK-0354, or (B) monomethyl fumarate (MMF) for 10 minutes prior to measurement of activity using a BRET-based biosensor. Data is normalized to the percent maximum activity of nicotinic acid. EC50, E_min_, and E_max_ with corresponding statistics can be found in Supplementary Table 11. (C-D) β-arrestin 2 and GRK2 activity of HEK 293 cells transiently transfected with *HCAR2-*WT or *HCAR2-*p.R311C. Cells were stimulated with (C) MK-0354, or (D) MMF for 10 minutes prior to measurement of activity using a BRET-based biosensor. Data is normalized to the percent maximum activity of nicotinic acid EC50, E_min_, and E_max_ with corresponding statistics can be found in Supplementary Table 12. (E-F) Early endosome translocation activity of HEK 293 cells transiently transfected with *HCAR2-*WT or *HCAR2-*p.R311C. Cells were stimulated with (E) MK-0354, or (F) MMF for 30 minutes prior to measurement of activity using a BRET-based biosensor. Data is normalized to the percent maximum activity of nicotinic acid EC50, E_min_, and E_max_ with corresponding statistics can be found in Supplementary Table 13. (G) Immunoblots for HCAR2 and Hibit protein in whole cell lysates from CHO cells stably expressing a doxycycline (dox)-inducible Hibit-HCAR2 WT or p.R311C construct. Cells were treated with vehicle or dox overnight to induce expression. β-actin serves as the loading control. . For A-L, all experiments have been performed 3 separate times, and the average of 3 experiments is shown. G has been performed once as a secondary validation of Hibit results. All values presented as mean ± SEM.
